# Blood bacterial DNA, intestinal adenoma and colorectal cancer

**DOI:** 10.1101/2021.07.22.21260498

**Authors:** Massimiliano Mutignani, Roberto Penagini, Giorgio Gargari, Simone Guglielmetti, Marcello Cintolo, Aldo Airoldi, Pierfrancesco Leone, Pietro Carnevali, Clorinda Ciafardini, Giulio Petrocelli, Federica Mascaretti, Barbara Oreggia, Lorenzo Dioscoridi, Federica Cavalcoli, Massimo Primignani, Francesco Pugliese, Paola Bertuccio, Pietro Soru, Carmelo Magistro, Giovanni Ferrari, Michela Speciani, Giulia Bonato, Marta Bini, Paolo Cantù, Flavio Caprioli, Marcello Vangeli, Edoardo Forti, Stefano Mazza, Giulia Tosetti, Rossella Bonzi, Maurizio Vecchi, Carlo La Vecchia, Marta Rossi

## Abstract

**Objective:** We aimed to investigate the relation of blood bacterial DNA load and profiling with intestinal adenoma (IA) and colorectal cancer (CRC) patients.

**Design:** We performed 16S rRNA gene analysis of blood from 100 incident histologically confirmed CRC cases, 100 IA and 100 healthy subjects, matched to cases by centre, sex and age. Bacterial load was analysed using multiple conditional logistic regression. Differences in terms of abundance of bacteria between groups were estimated through analysis based on negative binomial distribution normalization. Random Forest was applied to predict the group assignment.

**Results:** We found an overrepresentation of blood 16S rRNA gene copies in colon cancer as compared to tumor-free controls (IA and healthy subjects). The odds ratio of colon cancer for the highest versus the lowest three quintiles of gene copies was 2.62. (95% confidence interval=1.22-5.65). No difference was found for rectal cancer and IA. For high 16S rRNA, community diversity was higher in colon cancers than controls. CRC cases had an enrichment of Peptostreptococcaceae and Acetobacteriaceae and a reduced abundance of Bacteroidaceae, Lachnospiraceae, and Ruminococcaceae. Identified variables predicted CRC from control and IA patients with an accuracy of 0.70.

**Conclusion:** Colon cancer patients had a higher DNA bacterial load and a different bacterial profiling as compared to healthy subjects, IA and rectal cancers, indicating a higher passage of bacteria from gastrointestinal tract to bloodstream. Further studies are needed to confirm this result and exploit it to conceive new non-invasive techniques for an early diagnosis of CRC.

## INTRODUCTION

Colorectal cancer (CRC) is the 3^rd^ more common cancer, and ranks second in terms of mortality, worldwide [1]. Although mortality trends have been favourable in Europe during the last decades, with a rate of 15.4/100 000 in men and 8.6/100 000 in women in 2020, in most eastern European countries CRC mortality is still increasing [2, 3, 4].

CRC derives from a sequential accumulation of genetic alterations that involves the transition from normal mucosa to pre-malignant lesions with progression to intestinal adenoma (IA) and invasive CRC [5]. Inflammation and immunity are inextricably linked to all phases of CRC development [6]. Numerous studies identified chronic intestinal inflammation as a risk factor for CRC, as also confirmed by the increase of the incidence of this tumour in inflammatory bowel disease patients [7, 8]. IA and CRC have also been associated with increased circulating inflammation [9, 10, 11, 12, 13], and more recently with dysfunction of the gut mucosal barrier [14]. The community structure of the intestinal microbial ecosystem influences the risk of IA and CRC [14, 15, 16, 17]. There is evidence that in gut mucosal microbiota *Fusobacterium* spp. were increased in CRC patients [15, 16, 17]; *Bacteroides fragilis* and the genus *Porphyromonas* have been also associated with an increased risk of CRC [16, 17], while the increase in members of the genus *Escherichia* was associated with a higher IA risk [17]. Numerous studies analyzed fecal microbiome, not always reporting consistent results [15, 18, 19, 20], but recent meta-analyses including geographically and technically different shotgun metagenomic studies showed higher fecal microbiota richness in CRC cases as compared to controls [21], and identified a set of bacterial taxa significantly enriched in CRC cases [21, 22, 23].

Increasing evidence has pointed out the presence of bacterial DNA in blood [24, 25]. Microbial signatures have been reported for gut dysbiosis among diabetic subjects and for liver fibrosis in obese patients [26, 27]. Microbiome analysis of blood has also been proposed as a tool to discriminate between cancer patients and healthy subjects [28]. In particular, differences in circulating bacterial factors can occur in CRC patients, in whom epithelial barrier dysfunction can lead to increased intestinal permeability, plausibly resulting into a greater bacterial translocation from the gastrointestinal tract to bloodstream in IA and/or CRC. Some differences in relative abundance of the bacterial DNA in plasma between healthy, IA and CRC subjects have been reported in a case-control study from China, including 57 participants, but differences in terms of total bacterial load have not been analyzed yet [29].

In this study, we aimed to compare the load of bacterial DNA in blood and taxonomic profile between CRC, IA and healthy controls, using data from an *ad hoc* developed study.

## MATERIALS AND METHODS

We conducted an observational study between May 2017 and November 2019 in the metropolitan area of Milan, Italy. Recruitment centres included two general hospitals of Milan: the Digestive and Interventional Endoscopy Unit, Azienda Socio Sanitaria Territoriale (ASST) Grande Ospedale Metropolitano Niguarda, the coordinator centre, and the Gastroenterology and Endoscopy Unit, Fondazione Istituto di Ricovero e Cura a Carattere Scientifico (IRCCS) Ca’ Granda Ospedale Maggiore Policlinico. Both hospitals included a colonoscopy screening referral centre of the CRC screening program, managed by Health Protection Agency.

CRC cases were enrolled together with non-cancer adenomatous polyps and healthy controls, frequency-matched with cases by center, age ± 5 years and sex. Trained interviewers recruited study participants among eligible outpatients or inpatients who were scheduled for a colonoscopy, including patients referred for the CRC screening program. Excluded criteria were: 1) colonoscopy in the last 5 days; 2) reported previous cancers; 3) inflammatory chronic bowel diseases, 4) liver or kidney failure (creatinine ≥1,7 mg/dl, dialysis); 5) NYHA grade III or IV heart failure; 6) primary or secondary immunodeficiency; 7) recent hospitalization (1 month) for immune, inflammatory, autoimmune diseases, or bacterial/viral infections, 8) blood transfusions in the previous year; 9) celiac disease and a relevant diet modification during the last month. IA and control subjects with previous colonoscopy/sigmoidoscopy with endoscopic resection of a colonic lesion were also excluded.

A total of 620 patients were contacted by the trained interviewers. Of these, around 25% did not meet the eligibility criteria and less than 2% refused to participate in the study. Furthermore, 49 subjects were excluded due to some inaccuracies in the enrolment procedures and 42 due to previous cancers or to other ineligible conditions that were discovered after further data check. From the remaining 347 patients, the final sample after matching included 300 subjects: 100 CRC cases, 100 IA patients and 100 healthy controls.

Colonoscopy and histological examinations were revised by two pathologists who determined CRC cases and their clinical characteristics (e.g. stage, lymph node), as well as IA and their major features (e.g. morphology, measure), and healthy controls.

Cases included 100 incident, histologically confirmed CRC: 62 men and 38 women (mean age: 67, range 31–85 years). Of these, 21 were in the right colon (International Classification of Diseases, 10th Edition, ICD-10, C18.0, C18.2, C18.3), 12 in the transverse colon, in the splenic flexure, and in the descending colon (ICD-10, C18.4, C18.5, C18.6), 17 in the sigmoid colon (ICD-10, C18.7), and 50 in the rectum, including the rectosigmoid junction (ICD-10, C20, C19.9).

One hundred IA patients (mean age: 66, range 34–84 years) and 100 healthy controls (mean age: 66, range 26–85 years) were included.

The study protocol was revised and approved by the ethical committees of the hospitals involved in data recruitment: ASST Grande Ospedale Metropolitano Niguarda (No. 477-112016) and Fondazione IRCCS Ca’ Granda Ospedale Maggiore Policlinico (No. 742-2017).

### Interview

After written consent, a *face to face* interview was performed. The questionnaire included information on socio-demographics, smoking habits, physical activity, anthropometric measures, occupational exposures, medical history, selected drug and supplement use, family history of cancer, sleeping habits and dental care. A food frequency questionnaire (FFQ), based on an Italian reproducible and valid FFQ [30, 31], was used to assess the past patients’ usual diet.

### Blood collection

Blood samples were collected before the colonoscopy in order to avoid possible bacteria contamination after the colonscope insertion and to keep the same setting for each participant. An aliquot of 7 ml of blood was collected in a tube with EDTA and an aliquot of 3 ml in a blank (without anticoagulant) tube. Three microvials of 1 ml from EDTA tube were immediately stored at −80 ºC for the microbiomic analysis. The remaining blood was processed and centrifuged, and then stored at −80º C. At the end of data recruitment, blood samples were sent to Vaiomer SAS, Labège, France, for the analysis of the microbiome. To avoid the possibility that differences between groups could be due to experimental biases, the operators were blind to the group assignment and the samples were analyzed in the same experiment, with the same reagent batches and manipulator, in order to keep the signal to noise ratio optimal and to reduce technical variability.

### DNA extraction, qPCR experiments and sequencing of 16S rRNA gene amplicons

Bacterial DNA quantification and sequencing reactions were performed by Vaiomer SAS using an optimized blood-specific technique [24, 32, 33]. DNA was extracted from 0.25 ml of whole blood and collected in a final 50 μl extraction volume. Real-time polymerase chain reaction (PCR) amplification was performed using panbacterial primers EUBF 5’-TCCTACGGGAGGCAGCAGT-3’ and EUBR 5’-GGACTACCAGGGTATCTAATCCTGTT-3’ [34], which target the V3-V4 hypervariable regions of the bacterial 16S rRNA gene with 100% specificity (i.e., no eukaryotic, mitochondrial, or Archaea DNA is targeted) and high sensitivity (16S rRNA of more than 95% of bacteria in Ribosomal Database Project database are amplified). The abundance of the 16S rRNA gene in blood samples was measured by qPCR in triplicate and normalized using a plasmid-based standard range. The results were reported as number of copies of 16S rRNA gene per µl of blood. DNA from whole blood was also used for 16S rRNA gene taxonomic profiling using MiSeq Illumina technology using the 2 x 300 paired-end MiSeq kit V3. The samples 20056, 10251, 10248 and 20086 (referring to 2 IA and 2 control subjects) were excluded from the diversity analyses as they did not reach the threshold of 5,000 reads.

Then, sequences were analyzed using Vaiomer bioinformatic pipeline to determine bacterial community profiles. Briefly, after demultiplexing of the bar-coded Illumina paired reads, single read sequences were trimmed (reads R1 and R2 to respectively 290 and 240 bases) and paired for each sample independently into longer fragments, non-specific amplicons were removed and remaining sequences were clustered into operational taxonomic units (OTUs) using FROGS v1.4.0 [35] with default parameters. A taxonomic assignment was performed by Blast+ v2.2.30 against the Silva 132 Parc database. The OTUs were clustered based on 97% sequence similarities by two steps through swarm algorithm v2.1.6. The first step consisted of a clustering with an aggregation distance equal to 1. The second step consisted of a clustering with an aggregation distance equal to 3. OTUs with relative abundance lower than 0.005% of the whole dataset of reads were removed. All the reads are publicly available in the European Nucleotide Archive (ENA) with the accession number: PRJEB46474.

### Bacterial DNA contamination assessment

To assess the potential bacterial DNA contamination from environment and reagents, several negative controls were included to the analyses (Methods Supplementary). This analysis showed that the background noise and blood contamination did not impact the results of this study.

### Statistical analyses

Two-tailed Wilcoxon signed-rank tests and Friedman tests were used to compare 16S rRNA gene between groups. Odds ratios (OR) of CRC cases and their corresponding 95% confidence intervals (CI) were estimated as compared to control and IA subjects through logistic regression models conditioned on the matching variable. The number of 16S rRNA gene copies was included in the models as quintiles (categorically) based on the distribution of control and IA subjects, and as continuous variables, with the measurement unit sets to the difference between the upper cut-points of the 4th and 1st quintile, equal to 4328. Tests for trend were based on the likelihood ratio test between models with and without a linear term. Multinomial logistic regression was used to estimate separate ORs for colon and rectal cancer and to test for heterogeneity between the two sites.

Analysis on alpha-diversity and beta-diversity indices, as well as other taxonomic variables were computed among 296 subjects (because of 4 missing data due to technical reasons described above). To assess the samples diversity in terms of richness and evenness, various alpha-diversity indices, including Observed, Chao1, Shannon, Simpson and InvSimpson, were calculated by R PhyloSeq v1.14.0 package. Two-tailed Mann-Whitney tests were used to determine differences in terms of alpha-diversity between groups.

To estimate the beta-diversity, Permutational Multivariate Analysis of Variance Suing Distance Matrices (PERMANOVA) was applied based on the UniFrac distances, and Principal Coordinates Analysis (PCoA) was applied to visualize possible differences between groups.

Differences in terms of bacterial taxa and OTUs were evaluated through Welch test after DESeq2 normalization of data, based on negative binomial distribution (R package “DESeq2” v1.26.0).

For each statistical analysis, a post-hoc p-value adjustment was performed using the Hochberg-Benjamin correction, when appropriate.

Random Forest (R libraries: “randomforest”, “caret”, “Boruta”) was used to infer whether there was a set of variables that were able to discriminate which group the samples belong to. For supervised methods, in order to decrease the background noise due to the different library size of the samples sequenced, the data were normalized: the relative abundance of taxa was multiplied by 16S rRNA gene abundance (determined by qPCR) in each blood sample.

## RESULTS

Table 1 gives the distribution of 100 healthy controls, 100 IA patients, and 100 cases of CRC according to sex, age, study centre and education. By design, the three groups had the same sex and centre distributions and were similar in terms of age. Cases tended to be less educated than IA subjects and controls in absence, however, of a significant difference (χ^2^ test p=0.18).

**TABLE 1.** Distribution of 100 healthy controls, 100 intestinal adenoma (IA) patients and 100 colorectal cancer (CRC) cases by sex, age, study center and years of education. Italy 2017-2019.

| Characteristic | Controls | IA | CRC |
| --- | --- | --- | --- |
| Sex |  |  |  |
| Male | 62 (62%) | 62 (62%) | 62 (62%) |
| Female | 38 (38%) | 38 (38%) | 38 (38%) |
| Age group (yr) |  |  |  |
| <50 | 7 (7%) | 4 (4%) | 10 (10%) |
| 50-59 | 23 (23%) | 20 (20%) | 19 (19%) |
| 60-69 | 26 (26%) | 36 (36%) | 29 (29%) |
| 70-79 | 33 (33%) | 29 (29%) | 31 (42%) |
| ≥80 | 11 (11%) | 11 (11%) | 11 (11%) |
| $\chi^2$ test, p=0.76 | | | |
| Mean (SD) age (years) * | 66.0 (11.8) | 65.9 (10.9) | 66.1 (11.6) |
| Centre |  |  |  |
| Niguarda | 65 (65%) | 65 (65%) | 65 (65%) |
| Policlinico | 35 (35%) | 35 (35%) | 35 (35%) |
| Education (yr) |  |  |  |
| <7 | 12 (12%) | 19 (19%) | 25 (25%) |
| 7-11 | 24 (24%) | 25 (25%) | 25 (25%) |
| ≥12 | 64 (64%) | 56 (56%) | 50 (50%) |
| $\chi^2$ test, p=0.18 | | | |

\* Anova test for heterogeneity p=1.00. SD: standard deviation.

### 16S rRNA gene copies

We found an overall mean of 7687 16S rRNA gene copies per µl of blood, with a mean of 7628 among controls, 7586 among IA and 8387 among CRC subjects (9145 in colon and 7629 in rectal cancers), with no significant differences between the three groups (p for heterogeneity=0.482) (Figure 1, Supplementary). Since 16S rRNA gene copy distribution was very similar in control and IA subjects (p for heterogeneity=0.95), we grouped them as reference group and compared their 16S rRNA gene copy distribution with that of CRC, colon and rectal cancer cases. We did not find heterogeneity between CRC and control/IA groups (p=0.336), whereas significant differences emerged between colon cancer and control/IA (p =0.025; Figure 1).

**Figure 1.**
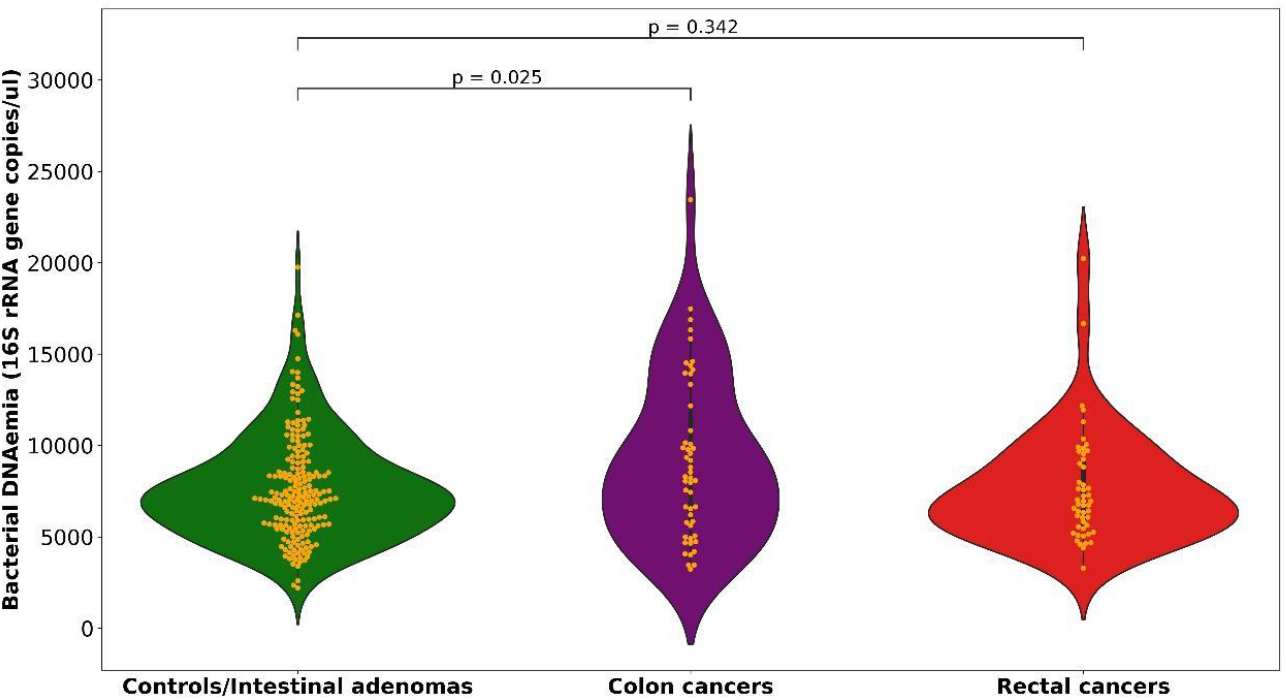
Distribution of 16S rRNA gene copies per µl of whole blood among controls/intestinal adenomas, colon and rectal cancers.

Table 2 shows the distribution of control/IA subjects, CRC, colon and rectal cancer cases, the ORs and the corresponding 95% CIs according to quintiles of 16S rRNA gene copies, as well as the continuous OR for an increment of around 4300 gene copies. We found a direct association of 16S rRNA gene copies with colon cancer. Subjects in the highest quintile of gene copies (≥ 9707copies) had an OR of 2.62 for colon cancer as compared to those in the first three quintiles (<7618 copies). The association significantly increased after the fourth quintile (p for trend=0.013) and became stronger for levels higher than the fifth quintile cut-off. The OR was 7.22 (95%CI= 2.18-23.9) for the 90^th^ centile (>11 265 copies) and 17.08 (95%CI= 3.36-86.87) for the 95^th^ centile (>13 000 copies) as compared to the lowest three quintiles. In addition, the continuous OR indicated a two-fold increased risk for an increment equal to 4328 copies (OR=2.02; 95%CI=1.26-3.25).

**Table 2.**
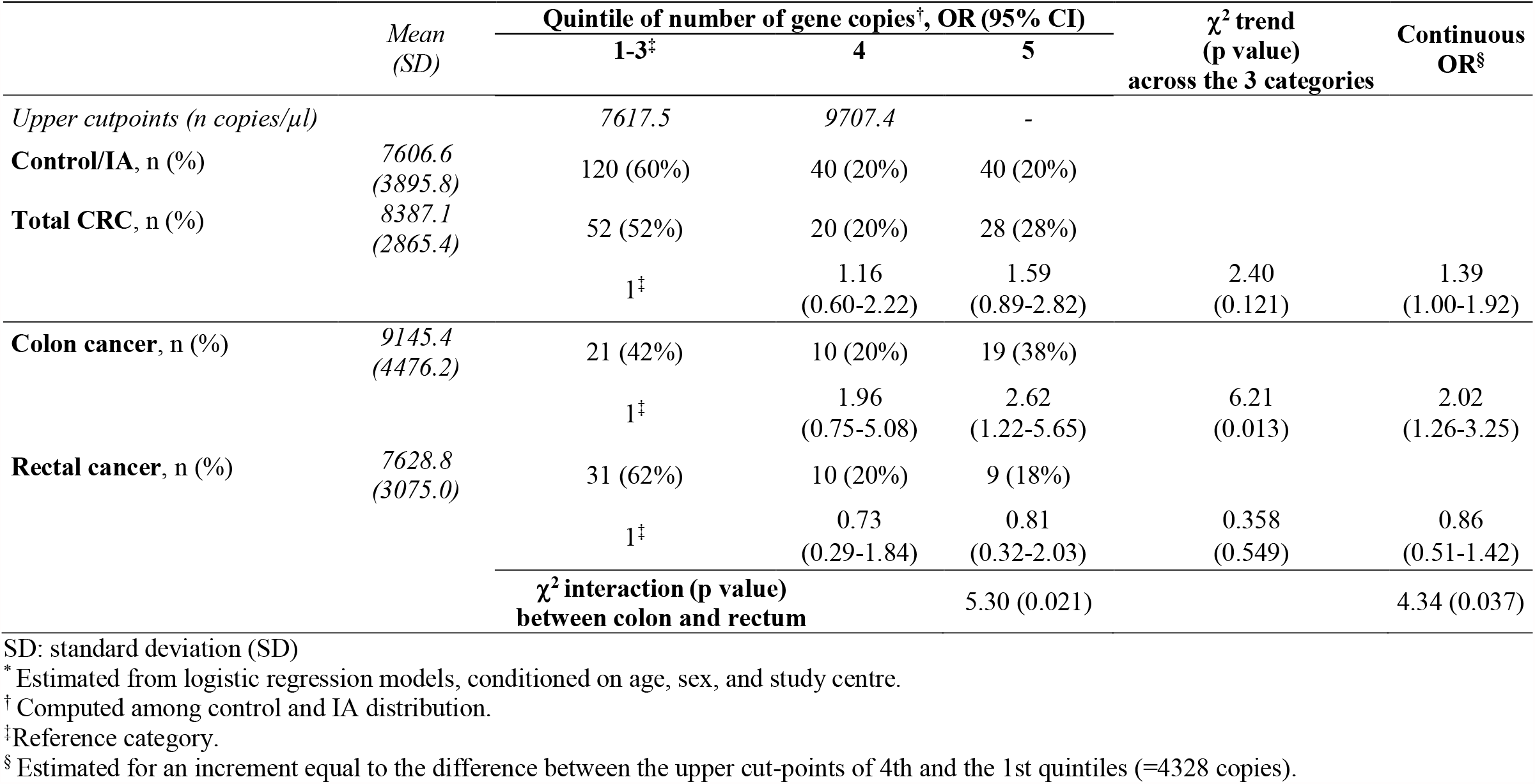
Odds ratios (OR)^*^ and corresponding 95% confidence intervals (CI) according to quintiles of 16S rRNA gene copies in whole blood/µl of 100 control and 100 intestinal adenoma (IA) patients and of 100 colorectal cancer (CRC) cases (50 colon and 50 rectal cancers). Italy 2017-2019.

In contrast, no association was found between 16S rRNA gene copies and rectal cancer. The OR for the highest versus the lowest three quintiles was 0.81 (95%CI=0.32-2.03) and the continuous OR was 0.86 (95%CI=0.51-1.42). The heterogeneity between colon and rectal was significant across quintiles (p=0.021) and in continuous (p=0.037).

The OR of CRC for the highest versus the lowest three quintiles was 1.59 (95%CI=0.89-2.82) and the continuous OR was 1.39 (95%CI=1.00-1.92).

When the association with 16S rRNA gene copies was further examined according to the colon subsites, the positive association appeared to be more pronounced for right colon, with an OR for the highest versus the lowest three quintiles of 10.75 (95%CI=2.16-53.42) as compared to 1.25 (95%CI=0.46-3.38) for other colon sites, in absence, however, of heterogeneity (p=0.123). Among 21 right colon cancers, 11 (53%) were in the highest quintile of gene copies, whereas among 29 other colon cancer subsites, 8 (26%) were in the highest quintile, in comparison to 40 out of 200 (20%) among control/IA subjects. In particular, among 11 cancers of the ascending colon, 7 (64%) were in the highest quintile (Table 3).

**Table 3.** Distribution of control and intestinal adenoma (IA) subjects, and colon and rectal cancer cases by cancer subsites and quintiles of 16S rRNA gene copies.

|  | Total | Quintile of number of gene copies*, n (%) |  |  |
| --- | --- | --- | --- | --- |
|  |  | 1-3 | 4 | 5 |
| *Control/IA* | 200 | 120 (60%) | 40 (20%) | 40 (20%) |
| Right colon | 21 | 7 (33%) | 3 (14%) | 11 (53%) |
| Cecum | 4 | 2 (50%) | 0 (0%) | 2 (50%) |
| Ascending | 11 | 2 (18%) | 2 (18%) | 7 (64%) |
| Hepatic flexure | 6 | 3 (50%) | 1 (17%) | 2 (33%) |
| Other than right colon | 29 | 14 (48%) | 7 (24%) | 8 (38%) |
| Transverse colon | 2 | 1 (50%) | 1 (50%) | 0 (0%) |
| Splenic flexure | 3 | 1 (33%) | 1 (33%) | 1 (33%) |
| Descending colon | 7 | 4 (57%) | 2 (29%) | 1 (14%) |
| Sigmoid colon | 17 | 8 (47%) | 3 (18%) | 6 (35%) |
| Rectum | 50 | 31 (62%) | 10 (20%) | 9 (18%) |
| Rectosigmoid junction | 3 | 3 (100%) | 0 (0%) | 0 (0%) |
| Rectum | 47 | 28 (60%) | 10 (21%) | 9 (19%) |

\* Computed among control and IA distribution.

### Alpha and beta diversity

No differences between groups were found in terms of α-diversity indices (Supplementary Table 1). When we restricted the analyses to the subjects in the two highest quintiles of 16S rRNA gene copies, we found a higher diversity in colon cancer cases as compared to controls in terms of Observed taxa and Chao1 indices for both genera (median of 32 vs 28, p=0.054 and median of 49 vs 40.6, p=0.059, respectively) and OTUs (median of 40 vs 34, p=0.039 and median of 71.1 vs 53.4, p=0.067), and as compared to rectal cancer cases in terms of both Observed genera and OTUs (median of 32 vs 29, p=0.023 and median of 40 vs 37, p=0.029) (Table 4, Figure 2). Colon cancers also appeared to be higher than IA in terms of Observed genera (median of 32 vs 28, p=0.071), and when we compared the Observed genera index in colon cancer cases versus control/IA subjects together, the p value for heterogeneity decreased to 0.035.

**Table 4.** Distributions of Observed, Chao1, Shannon and Simpson alpha-diversity indices of control, intestinal adenoma (IA), colon and rectal cancer subjects for bacterial genera and OTUs among the highest two quintiles of 16S rRNA gene copies.

|  |  | Median<br>(I-III quartiles) |  |  |  | p* | p* | p* | p* | p* | p* |
| --- | --- | --- | --- | --- | --- | --- | --- | --- | --- | --- | --- |
| *Alpha-diversity* |  | Controls | IA | Colon cancer | Rectal cancer | Controls vs IA | Colon cancers vs IA | Colon cancer vs controls | Rectal cancer vs IA | Rectal cancer vs controls | Colon cancer vs rectal cancer |
| **Genera** | *Observed* | 28<br>(25-31) | 28<br>(25-33.5) | 32<br>(26-39) | 29<br>(25-31) | 0.942 | 0.071 | 0.054 | 0.714 | 0.968 | 0.023 |
|  | *Chao1* | 40.6<br>(31.7-52.3) | 42.5<br>(33.4-52.5) | 49<br>(41.5-59) | 41<br>(35-46) | 0.476 | 0.148 | 0.059 | 0.703 | 0.789 | 0.080 |
|  | *Shannon* | 2.33<br>(2.05-2.55) | 2.41<br>(2.18-2.56) | 2.33<br>(2.08-2.58) | 2.26<br>(2.06-2.44) | 0.254 | 0.535 | 0.614 | 0.119 | 0.715 | 0.442 |
|  | *Simpson* | 0.86<br>(0.80-0.89) | 0.87<br>(0.84-0.89) | 0.84<br>(0.81-0.89) | 0.86<br>(0.79-0.88) | 0.394 | 0.496 | 0.846 | 0.186 | 0.697 | 0.513 |
| **OTUs** | *Observed* | 34<br>(30-39) | 35<br>(32.4-43.5) | 40<br>(33-51) | 37<br>(30-38) | 0.413 | 0.154 | 0.039 | 0.570 | 0.820 | 0.029 |
|  | *Chao1* | 53.4<br>(47.8-70.5) | 66<br>(51.9-92.8) | 71.1<br>(52-87) | 56<br>(51-73) | 0.070 | 0.981 | 0.067 | 0.233 | 0.565 | 0.278 |
|  | *Shannon* | 2.52<br>(2.27-2.74) | 2.63<br>(2.46-2.84) | 2.60<br>(2.46-2.73) | 2.50<br>(2.27-2.65) | 0.154 | 0.662 | 0.473 | 0.089 | 0.727 | 0.149 |
|  | *Simpson* | 0.90<br>(0.86-0.93) | 0.91<br>(0.88-0.93) | 0.90<br>(0.87-0.92) | 0.87<br>(0.86-0.91) | 0.233 | 0.473 | 0.653 | 0.062 | 0.403 | 0.229 |

\* P for heterogeneity estimated from the Wilcoxon rank-sum

**Figure 2 A.**
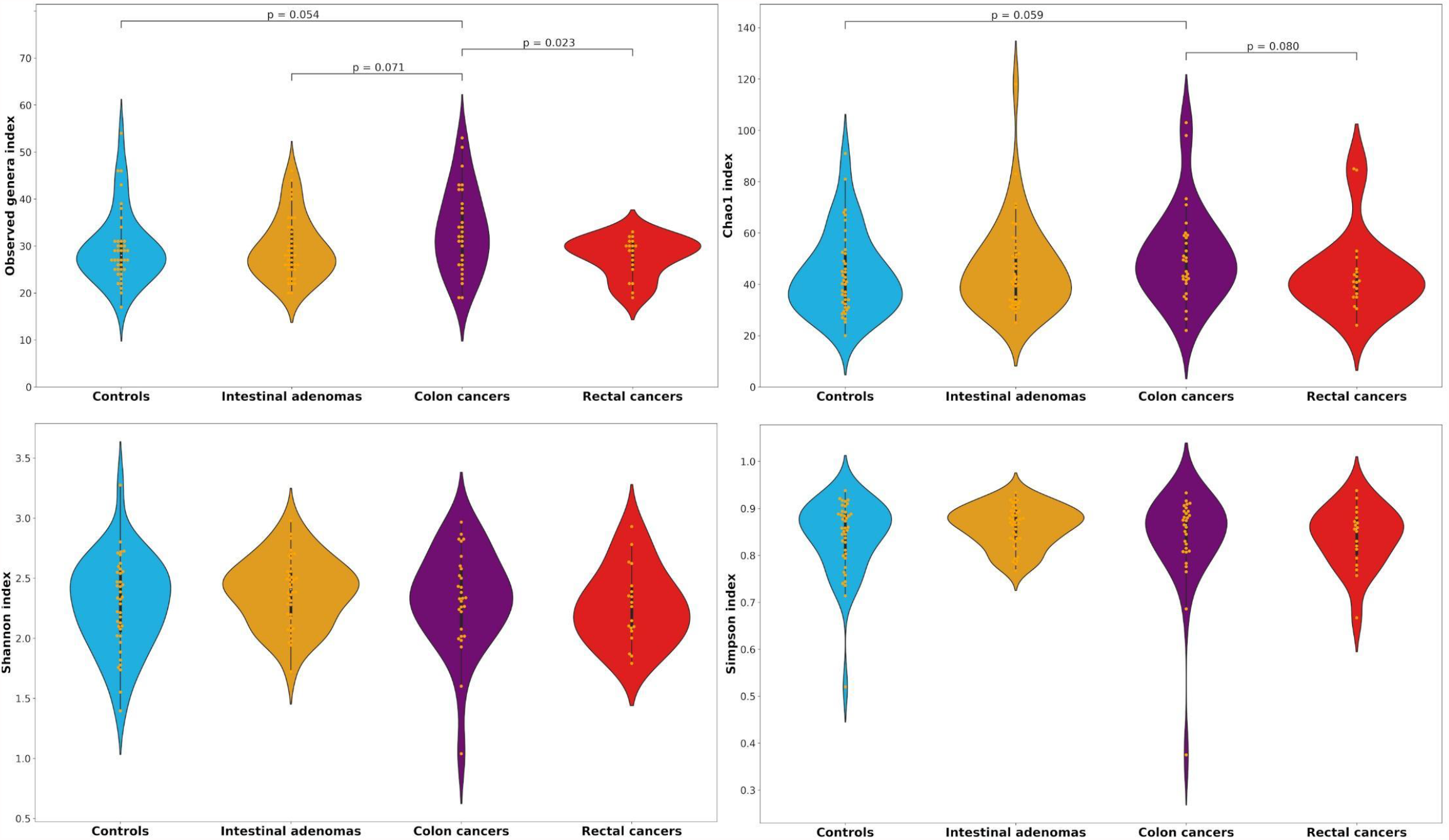
Distributions of Observed, Chao1, Shannon and Simpson alpha-diversity indices among controls, intestinal adenomas, colon and rectal cancers for genera among the highest two quintiles of 16S rRNA gene copies.

**Figure 2 B.**
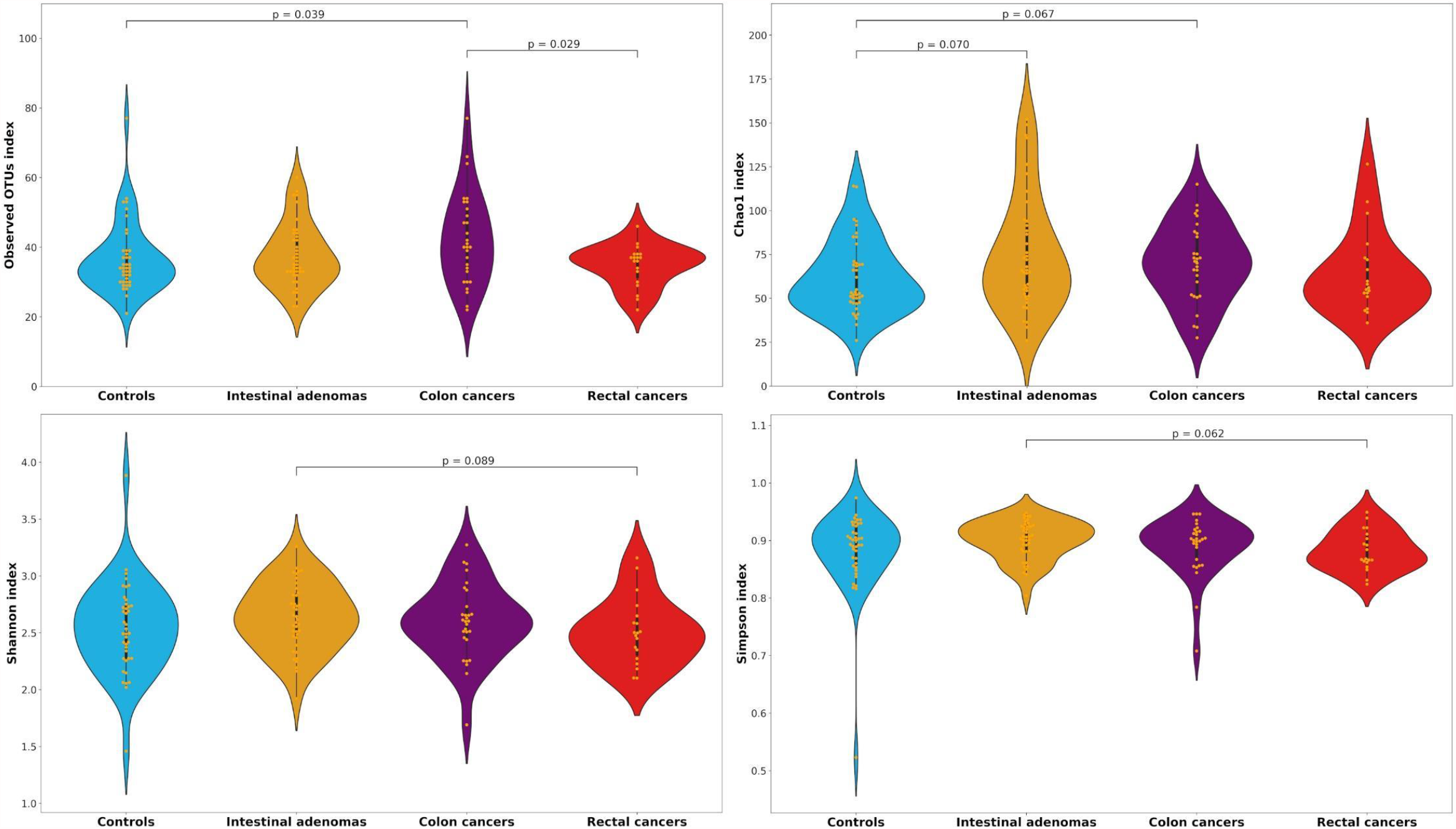
Distributions of Observed, Chao1, Shannon and Simpson alpha-diversity indices among controls, intestinal adenomas, colon and rectal cancers for the operational taxonomic units (OTUs) among the highest two quintiles of 16S rRNA gene copies.

Concerning the beta diversity, no differences between groups were found overall and among subjects into the two highest quintiles of 16S rRNA gene copies. However, when we further restricted the analyses to the highest quintile of 16S rRNA, we observed significant differences between control/IA, colon and rectal cancer patients (Weighted UniFrac, p= 0.026; Unweighted UniFrac, p=0.051; Generalized UniFrac, p=0.031) (Figure 3 – A, B, C). Post-hoc analyses splitting the groups 2 by 2 showed a trend between controls/IA and colon cancers in Weighted UniFrac (p=0.073) (Figure 3 – D).

**Figure 3.**
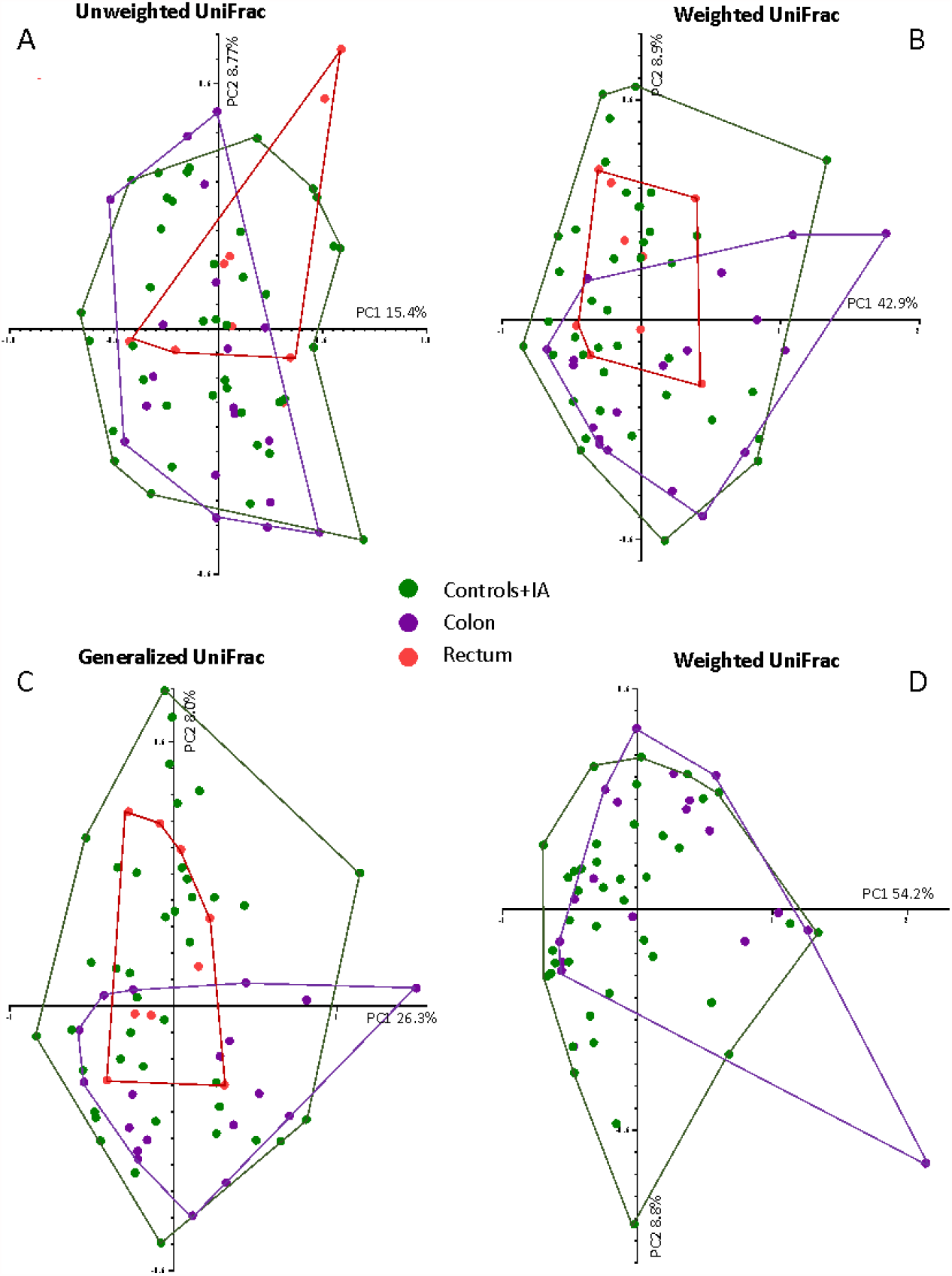
®-diversity of controls and intestinal adenoma (Controls+IA) group, colon cancer (Colon) and rectal cancer (Rectum) among subjects into the fifth quintile of 16S rRNA gene copies. The figure shows the UniFrac ®-diversity in all the three variants: (A) Unweighted UniFrac; (B) Weighted UniFrac; (C) generalized UniFrac for all the three groups: Controls+IA, Colon and Rectum. Panel (D) shows the Weighted Unifrac for Controls+IA and colon cancers after post-hoc analyses.

### Taxonomic profiling of blood bacterial DNA between groups

We detected a total of 1081 OTUs that were taxonomically classified into 15 phyla, 34 classes, 87 orders, 164 families and 325 genera.

Pseudomonadaceae, Micrococcaceae, Burkholderiaceae, Caulobacteraceae, Moraxellaceae and Flavobacteriaceae were the six most represented families, which together accounted for more than 50% of all reads assigned to bacterial taxa (Supplementary Figure 2).

The mean of DESeq2 normalized data and the adjusted p values from the Welch test comparing every two groups (CRC versus control, CRC versus IA and IA versus control) and CRC versus control/IA on all the taxonomy levels and OTUs are shown in Supplementary Figure 3 and Figure 4, respectively. Several taxa differed between the groups. In particular, CRC samples were characterized by the increase of sequencing reads assigned to the bacterial families Peptostreptococcaceae and Acetobacteriaceae, together with a lower representation of the bacterial families Bacteroidaceae, Lachnospiraceae, and Ruminococcaceae (Figure 4).

**Figure 4.**
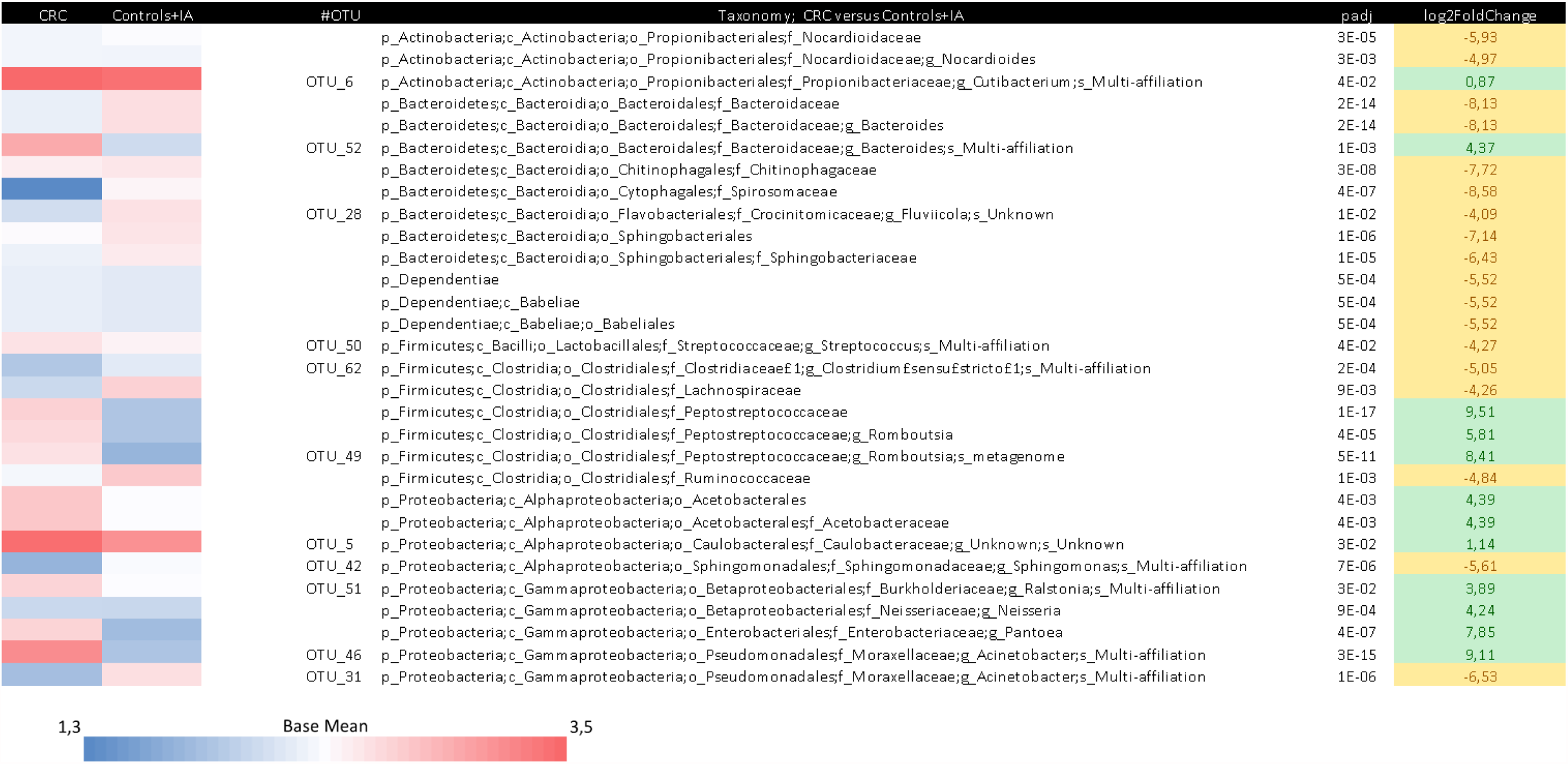
Different taxa between colorectal cancer (CRC) cases and controls and intestinal adenomas (IA) group by DESeq2 analyses. The taxonomic lineage of each taxon is shown: p, phylum; c, class; o, order; f, family; g, genus; OTU#, Operational Taxonomic Unit. The first two columns show the logarithmic transformation of normalized base mean value for each group. The “padj” column shows the p-value for heterogeneity between groups adjusted for multi-testing analyses. Positive fold changes (shown on a green background) designate taxon overrepresentation in the CRC group. Negative fold changes (shown on a yellow background) designate taxon underrepresentation in the CRC group.

Through the Random Forest supervised method, we found a set of variables that predict a CRC case versus controls/IA subjects with an accuracy of 0.70 (Sensitivity = 0.45; Specificity = 0.87) and another model that inferred the location of CRC discriminating colon from rectal cancer with an accuracy of 0.77 (Sensitivity = 0.71; Specificity = 0.82) (Figure 5). The first model inferred that the families Acetobacteraceae, Peptostreptococcaceae and Oligoflexaceae, the genus *Melittangium*, and the OTUs belonging to the genera *Acinetobacter, Pelomonas, Novosphingobium* and *Pajaroellobacter* were the most important variables to predict between CRC or control/IA group (Figure 5-A). The biplot in Figure 5-A shows a separation between CRC and control/IA subjects due to a higher dispersion of CRC cases. The second model inferred that the families Peptosteptococcaceae, Streptococcaceae and Ruminococcaceae, the genera *Arthrobacter, Clostridium* sensu stricto and *Kocuria*, and the OTUs belonging to the genera *Legionella, Kocuria* and *Lepisosteus oculatus* were the most important variables to predict the group between colon and rectal cancer, together with other variables that contributed to increase the accuracy of the model, including 16S rRNA gene copies, the phylum Proteobacteria, the order Rhizobiales and the genus *Bacteroides* (Figure 5 – B).

**Figure 5.**
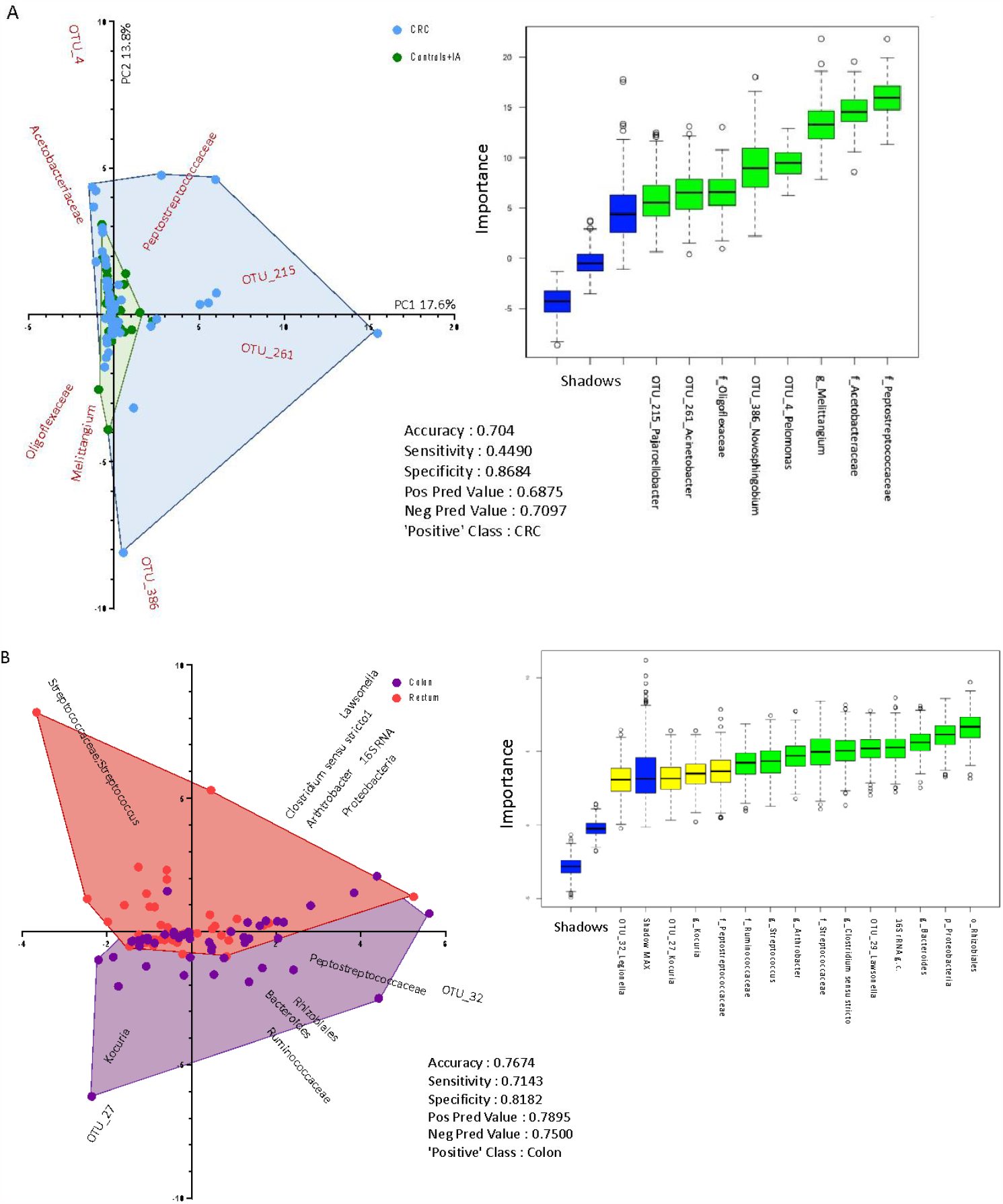
Biplot of predictive variables discriminating (A) colorectal cancer (CRC) versus control and intestinal adenoma (IA) subjects; (B) colon versus rectal cancer cases, using Random Forest algorithm. The boxplot on the right side of each figure shows the importance (based on mean accuracy level) of the variables by Boruta feature selection. The shadows are part of the Boruta algorithm and show the max, medium and lowest level of mean accuracy, using the same dataset with group labels shuffled. The table in the middle part of each figure shows the Random Forest results considering the ‘Positive’ Class as indicated.

When the Random Forest algorithm was applied to the sample belonging to the highest quintile of 16S rRNA gene copies, the accuracy to predict between CRC and control/IA group increased to 0.79.

## DISCUSSION

This study shows that colon cancer patients have an overrepresentation of bacterial DNA in blood as compared to tumor-free controls, including IA or healthy subjects. These results appeared stronger for cancers in the right colon, whereas no difference in terms of bacterial load was found for rectal cancer. For high levels of 16S rRNA gene copies (> 7618), colon cancers had increased community diversity but did not differ on community evenness from controls.

To our knowledge, no other study conducted an *ad hoc* data collection to systematically investigate blood microbiota in relation to CRC and/or IA to date. In a Chinese study, circulating bacterial DNA of 25 CRC, 10 IA and 22 healthy subjects was analysed through whole genome sequencing techniques on plasma samples [29], suggesting that the *Flavobacterium* DNA relative abundance was reduced in CRC and IA (<1%) as compared to control subjects (9.4%); on the contrary, there was a 10-fold increase DNA abundance of genus *Ruminococcus* in CRC (0.2%) as compared to controls (0.02%). In the same study, various attempts to identify bacterial biomarkers of CRC or IA through Random Forest algorithms were proposed, reporting a set of 28 species as important features to discriminate between CRC/IA group and controls, but results were based on small samples (23 and 34 subjects for discovery and validation cohort, respectively). Messaritakis et al. used PCR for the amplification of genomic DNA on blood in order to compare 397 adjuvant or metastatic CRC patients with 32 healthy controls in terms of the presence of 3 bacterial genes [36]. Significantly higher rates of glutamine synthase gene of *Bacteroides fragilis* and 5.8S rRNA of *Candida albicans* were observed in CRC patients (p < 0.001), especially in metastatic disease, suggesting a prognostic value of the detection of microbial translocation in blood. No association was found for the genus *E. coli*.

In our data, CRC patients had an enrichment of Peptostreptococcaceae and Acetobacteriaceae and a reduction of Bacteroidaceae, Lachnospiraceae, and Ruminococcaceae. The latter families are the most represented in the fecal and intestinal microbiota [37], while Peptostreptococcaceae and Acetobacteriaceae are less represented in the human intestine. These two families were found to be more abundant in chronic kidney disease (CKD) than healthy subjects in a case-control study on blood microbiome including 20 CKD cases and 20 controls [38]. In a study involving 99 CRC cases and 103 controls, high abundance of Lachnospiraceae was negatively correlated with colonic colonisation by oral bacteria, including oral pathogens associated with CRC, suggesting a protective role of Lachnospiraceae, potentially influenced by Western dietary patterns [39]. Ruminococcaceae abundance was found to decrease in CRC tissue as compared to tumor-adjacent biopsies and stool samples from the same case in a study including 294 subjects [40]. Moreover, the members of the families Lachnospiraceae and Ruminococcaceae were recognized as the most active members of the human colonic microbiota [41], able to efficiently convert fibres into butyrate [42], a bacterial catabolite widely demonstrated to regulate T-reg lymphocyte priming preventing CRC [43, 44].

Gut microbiota, inflammation and nutrition play an important role on intestinal permeability which may influence the risk of CRC [45, 46]. Bacteria have shown capacity to interact directly with immune system cells and to impact in multiple host functions [47, 48], but it is unclear which one comes first between local inflammation, intestinally permeability and changes in resident microbiota [49, 50]. In this context, it has also been shown that bacteria can disseminate to liver through the disruption of gut vascular barrier in colorectal cancer with hepatic metastasis [51].

Our data corroborate the hypothesis of a greater bacterial translocation from gastrointestinal tract to bloodstream in colon cancer, especially in right colon cancer, but not in rectal cancer patients. Along this line, various factors and biological aspects were different according to CRC sites. Physical activity, antibiotic use and family history of CRC were relevant in colon but not in rectal cancer [52, 53, 54, 55], and associations of some dietary components were found to be stronger for the cancer of some colon subsites [56, 57]. Moreover, the proximal and distal colon have a different embryological origin, also resulting in a distinct vascular supply [58]. Differences in underlying genetics, including genetic expression and immunological activity have been highlighted [59], with a negative gradient of immune cells from proximal colon to distal colon and rectum [58]. Gut microbiota and mechanisms of carcinogenesis also vary along the lower intestinal tract [54, 60, 61]. Supporting our hypothesis of a differential translocation, an important role may also be played by mucus layers, which were found to vary along the colon in mice in terms of O-glycosylated entities of Muc2 and of the host-microbiota symbiosis regulation [62].

Recent meta-analyses on faecal microbiome suggested universal, validated predictive taxonomic and functional microbiome CRC signatures, revealing potential mechanisms behind the intestinal carcinogenesis processes, and putting the basis for non-invasive CRC diagnosis through metagenomic analysis of faecal microbiome [21, 22, 23]. CRC screening programs can suffer from limited sensitivity and specificity of the tests used and from possible low adherence with CRC screening recommendations – mainly due to the refusal of faecal test and colonoscopy – in some countries. Innovative, non-invasive diagnostic tests would be of support for CRC control [63, 64]. Various tests based on blood analyses have been proposed, including techniques estimating the presence of *Streptococcus bovis* [65] or evaluating the antibody level against *Fusobacterium nucleatum* [66] in blood, and, more recently, the use of blood microbiome profile has been suggested [28].

Our data showed various taxa associated with CRC and identified a set of covariates of taxa and OTUs that is able to predict CRC from control/IA subjects and colon from rectal cancers with an accuracy around 0.7. When restricting the analyses to subjects with high levels of 16S rRNA gene copies, we found more accurate models, but we were not able to separate analysis for colon cancer only and results should be interpreted with caution given the small numbers. However, these findings can serve as a basis to conceive new non-invasive techniques for an early diagnosis of CRC based on bacterial DNA circulating in peripheral blood. In particular, they can be relevant for the detection of right colon cancers, which often have a subtle presentation and a more advanced stage at diagnosis, partly because right colon is more difficult to be explored, when compared to rectal and distal colon [67].

One of the strengths of this study is the conduction of an *ad-hoc* data collection, which includes a new developed standardized protocol, fully observed by the recruitment centres. CRC and the corresponding IA and control subjects were comparable in terms of setting since they derived from the same catchment area and recruitment procedures. Moreover, interviewers and investigators were blinded to the group assignment, as data collection was performed before endoscopy and diagnosis. Most cases were detected at the first CRC-diagnosing colonoscopy, allowing us to recruit truly incident cases, characterized by a minimal time between participant’s recruitment data and cancer diagnosis and with available clinical data from the very beginning of the diagnostic process. Moreover, the presence of healthy controls allowed a clean comparison with CRC and IA, and the inclusion of IA allowed to investigate an important phase on the mechanisms behind the process of the adenoma-carcinoma sequence. We were also able to adjust for study centre, sex and age, eliminating possible confounding effects of these covariates on 16s rRNA gene copies results.

In conclusion, our data confirm the presence of bacterial DNA in blood in healthy adults and indicate that colon cancer patients had a higher DNA bacterial load and a different bacterial profiling as compared to healthy, IA and rectal cancer subjects, revealing a higher passage of bacteria from gastrointestinal tract to bloodstream in colon cancer. Further studies are needed to confirm this result and possibly exploit it for the development of innovative early techniques for colon cancer diagnosis.

## Supporting information

Research checklist

## Data Availability

The epidemiological data that support the findings of this study are available upon reasonable request from the corresponding author (MR).
Raw metagenomic reads are publicly available in the European Nucleotide Archive (ENA) with the accession number: PRJEB46474.

## Acknowledgments

The authors would like to express their most sincere gratitude and appreciation to all participants and collaborators to this study, without whose effort this work would not have been feasible. A special thanks to Margherita Cozzi, Elena Tansi, Cinzia Della Noce, Rosa Restieri, Nadia Zaretti for their valuable involvement in this study. We thank all the nursing staff at the Digestive and Interventional Endoscopy Unit, ASST Grande Ospedale Metropolitano Niguarda, Milan, and at the Gastroenterology and Endoscopy Unit, Fondazione IRCCS Ca’ Granda Ospedale Maggiore Policlinico, Milan, and Valentina Taverniti, Guido Basilisco, Luca Elli, Francesca Ferretti, Gian Lorenzo Scacchi, Gian Eugenio Tontini, Giuseppe Torgano for their contribution in the data collection. A thankful mention to Annalisa Pascarella for the help in the graphic preparation and to Patrizia Riso for participating in the proof of concept of the project.

## Funding

This work was supported by the Italian Foundation for Cancer Research (AIRC) (My First AIRC grant no. 17070).

## Contributors

MM and RP contributed equally to this paper.

Conception and design: MR, SG, CLV, MP, RP, AA, MC, MauV and MM. Analysis of data: MR, GG and SG. Interpretation of data: MR, SG, CLV, RP, AA, MM, MC, GG and RB. Drafting the manuscript: MR, SG, GG, CLV, MS and RB. All authors contributed to data collection, and critical revision and final approval of the manuscript.

## Competing interests

The authors declare that there are no conflicts of interest

## Notes

### Competing Interest Statement

The authors have declared no competing interest.

### Author Declarations

The study protocol was revised and approved by the ethical committees of the hospitals involved in data recruitment: Azienda Socio Sanitaria Territoriale (ASST) Grande Ospedale Metropolitano Niguarda (No. 477-112016) and Fondazione Istituto di Ricovero e Cura a Carattere Scientifico (IRCCS) Ca' Granda Ospedale Maggiore Policlinico (No. 742-2017), Milan, Italy.

## BIBLIOGRAPHY

1 Sung H, Ferlay J, Siegel RL, Laversanne M, Soerjomataram I, Jemal A, et al. Global cancer statistics 2020. GLOBOCAN estimates of incidence and mortality worldwide for 36 cancers in 185 countries. CA Cancer J Clin 2021.

2 Carioli G, Bertuccio P, Boffetta P, Levi F, La Vecchia C, Negri E, et al. European cancer mortality predictions for the year 2020 with a focus on prostate cancer. Ann Oncol 2020;31:650–8.

3 Malvezzi M, Carioli G, Bertuccio P, Boffetta P, Levi F, La Vecchia C, et al. European cancer mortality predictions for the year 2018 with focus on colorectal cancer. Annals of oncology : official journal of the European Society for Medical Oncology 2018;29:1016–22.

4 Ait Ouakrim D, Pizot C, Boniol M, Malvezzi M, Boniol M, Negri E, et al. Trends in colorectal cancer mortality in Europe: retrospective analysis of the WHO mortality database. BMJ 2015;351:h4970.

5 Vogelstein B, Papadopoulos N, Velculescu VE, Zhou SB, Diaz LA, Kinzler KW. Cancer Genome Landscapes. Science 2013;339:1546–58.

6 Terzic J, Grivennikov S, Karin E, Karin M. Inflammation and colon cancer. Gastroenterology 2010;138:2101–14 e5.

7 Eaden JA, Abrams KR, Mayberry JF. The risk of colorectal cancer in ulcerative colitis: a meta-analysis. Gut 2001;48:526–35.

8 Herrinton LJ, Liu L, Levin TR, Allison JE, Lewis JD, Velayos F. Incidence and mortality of colorectal adenocarcinoma in persons with inflammatory bowel disease from 1998 to 2010. Gastroenterology 2012;143:382–9.

9 de Waal GM, de Villiers WJS, Forgan T, Roberts T, Pretorius E. Colorectal cancer is associated with increased circulating lipopolysaccharide, inflammation and hypercoagulability. Sci Rep 2020;10:8777.

10 Huang WY, Berndt SI, Shiels MS, Katki HA, Chaturvedi AK, Wentzensen N, et al. Circulating inflammation markers and colorectal adenoma risk. Carcinogenesis 2019;40:765–70.

11 Godos J, Biondi A, Galvano F, Basile F, Sciacca S, Giovannucci EL, et al. Markers of systemic inflammation and colorectal adenoma risk: Meta-analysis of observational studies. World J Gastroenterol 2017;23:1909–19.

12 Zhang Y, Yu X, Yu E, Wang N, Cai Q, Shuai Q, et al. Changes in gut microbiota and plasma inflammatory factors across the stages of colorectal tumorigenesis: a case-control study. BMC Microbiol 2018;18:92.

13 Ghuman S, Van Hemelrijck M, Garmo H, Holmberg L, Malmstrom H, Lambe M, et al. Serum inflammatory markers and colorectal cancer risk and survival. Br J Cancer 2017;116:1358–65.

14 Liu X, Cheng Y, Shao L, Ling Z. Alterations of the Predominant Fecal Microbiota and Disruption of the Gut Mucosal Barrier in Patients with Early-Stage Colorectal Cancer. Biomed Res Int 2020;2020:2948282.

15 Song M, Chan AT, Sun J. Influence of the Gut Microbiome, Diet, and Environment on Risk of Colorectal Cancer. Gastroenterology 2020;158:322–40.

16 Ahn J, Sinha R, Pei Z, Dominianni C, Wu J, Shi J, et al. Human gut microbiome and risk for colorectal cancer. J Natl Cancer Inst 2013;105:1907–11.

17 Xu K, Jiang B. Analysis of Mucosa-Associated Microbiota in Colorectal Cancer. Med Sci Monit 2017;23:4422–30.

18 Eklof V, Lofgren-Burstrom A, Zingmark C, Edin S, Larsson P, Karling P, et al. Cancer-associated fecal microbial markers in colorectal cancer detection. Int J Cancer 2017;141:2528–36.

19 Shah MS, DeSantis TZ, Weinmaier T, McMurdie PJ, Cope JL, Altrichter A, et al. Leveraging sequence-based faecal microbial community survey data to identify a composite biomarker for colorectal cancer. Gut 2018;67:882–91.

20 Yu J, Feng Q, Wong SH, Zhang D, Liang QY, Qin Y, et al. Metagenomic analysis of faecal microbiome as a tool towards targeted non-invasive biomarkers for colorectal cancer. Gut 2017;66:70–8.

21 Thomas AM, Manghi P, Asnicar F, Pasolli E, Armanini F, Zolfo M, et al. Metagenomic analysis of colorectal cancer datasets identifies cross-cohort microbial diagnostic signatures and a link with choline degradation. Nat Med 2019;25:667–78.

22 Wirbel J, Pyl PT, Kartal E, Zych K, Kashani A, Milanese A, et al. Meta-analysis of fecal metagenomes reveals global microbial signatures that are specific for colorectal cancer. Nat Med 2019;25:679–89.

23 Dai Z, Coker OO, Nakatsu G, Wu WKK, Zhao L, Chen Z, et al. Multi-cohort analysis of colorectal cancer metagenome identified altered bacteria across populations and universal bacterial markers. Microbiome 2018;6:70.

24 Paisse S, Valle C, Servant F, Courtney M, Burcelin R, Amar J, et al. Comprehensive description of blood microbiome from healthy donors assessed by 16S targeted metagenomic sequencing. Transfusion 2016;56:1138–47.

25 Castillo DJ, Rifkin RF, Cowan DA, Potgieter M. The Healthy Human Blood Microbiome: Fact or Fiction? Front Cell Infect Microbiol 2019;9:148.

26 Lelouvier B, Servant F, Paisse S, Brunet AC, Benyahya S, Serino M, et al. Changes in blood microbiota profiles associated with liver fibrosis in obese patients: A pilot analysis. Hepatology 2016;64:2015–27.

27 Sato J, Kanazawa A, Ikeda F, Yoshihara T, Goto H, Abe H, et al. Gut dysbiosis and detection of “live gut bacteria” in blood of Japanese patients with type 2 diabetes. Diabetes Care 2014;37:2343–50.

28 Poore GD, Kopylova E, Zhu Q, Carpenter C, Fraraccio S, Wandro S, et al. Microbiome analyses of blood and tissues suggest cancer diagnostic approach. Nature 2020;579:567–74.

29 Xiao Q, Lu W, Kong X, Shao YW, Hu Y, Wang A, et al. Alterations of circulating bacterial DNA in colorectal cancer and adenoma: A proof-of-concept study. Cancer Lett 2021;499:201–8.

30 Franceschi S, Negri E, Salvini S, Decarli A, Ferraroni M, Filiberti R, et al. Reproducibility of an Italian food frequency questionnaire for cancer studies: results for specific food items. European journal of cancer (Oxford, England : 1990) 1993;29A:2298–305.

31 Decarli A, Franceschi S, Ferraroni M, Gnagnarella P, Parpinel MT, La Vecchia C, et al. Validation of a food-frequency questionnaire to assess dietary intakes in cancer studies in Italy. Results for specific nutrients. Annals of epidemiology 1996;6:110–8.

32 Gargari G, Mantegazza G, Taverniti V, Del Bo C, Bernardi S, Andres-Lacueva C, et al. Bacterial DNAemia is associated with serum zonulin levels in older subjects. Sci Rep 2021;11:11054.

33 Lluch J, Servant F, Paisse S, Valle C, Valiere S, Kuchly C, et al. The Characterization of Novel Tissue Microbiota Using an Optimized 16S Metagenomic Sequencing Pipeline. PLoS One 2015;10:e0142334.

34 Nadkarni MA, Martin FE, Jacques NA, Hunter N. Determination of bacterial load by real-time PCR using a broad-range (universal) probe and primers set. Microbiology (Reading) 2002;148:257–66.

35 Escudie F, Auer L, Bernard M, Mariadassou M, Cauquil L, Vidal K, et al. FROGS: Find, Rapidly, OTUs with Galaxy Solution. Bioinformatics 2018;34:1287–94.

36 Messaritakis I, Vogiatzoglou K, Tsantaki K, Ntretaki A, Sfakianaki M, Koulouridi A, et al. The Prognostic Value of the Detection of Microbial Translocation in the Blood of Colorectal Cancer Patients. Cancers (Basel) 2020;12.

37 King CH, Desai H, Sylvetsky AC, LoTempio J, Ayanyan S, Carrie J, et al. Baseline human gut microbiota profile in healthy people and standard reporting template. PLoS One 2019;14:e0206484.

38 Shah NB, Allegretti AS, Nigwekar SU, Kalim S, Zhao S, Lelouvier B, et al. Blood Microbiome Profile in CKD : A Pilot Study. Clin J Am Soc Nephrol 2019;14:692–701.

39 Flemer B, Warren RD, Barrett MP, Cisek K, Das A, Jeffery IB, et al. The oral microbiota in colorectal cancer is distinctive and predictive. Gut 2018;67:1454–63.

40 Shah MS, DeSantis T, Yamal JM, Weir T, Ryan EP, Cope JL, et al. Re-purposing 16S rRNA gene sequence data from within case paired tumor biopsy and tumor-adjacent biopsy or fecal samples to identify microbial markers for colorectal cancer. PLoS One 2018;13:e0207002.

41 Peris-Bondia F, Latorre A, Artacho A, Moya A, D’Auria G. The active human gut microbiota differs from the total microbiota. PLoS One 2011;6:e22448.

42 Vital M, Karch A, Pieper DH. Colonic Butyrate-Producing Communities in Humans: an Overview Using Omics Data. mSystems 2017;2.

43 Singh N, Gurav A, Sivaprakasam S, Brady E, Padia R, Shi H, et al. Activation of Gpr109a, receptor for niacin and the commensal metabolite butyrate, suppresses colonic inflammation and carcinogenesis. Immunity 2014;40:128–39.

44 Venkataraman A, Sieber JR, Schmidt AW, Waldron C, Theis KR, Schmidt TM. Variable responses of human microbiomes to dietary supplementation with resistant starch. Microbiome 2016;4:33.

45 Bhat AA, Uppada S, Achkar IW, Hashem S, Yadav SK, Shanmugakonar M, et al. Tight Junction Proteins and Signaling Pathways in Cancer and Inflammation: A Functional Crosstalk. Front Physiol 2018;9:1942.

46 Yu LC. Microbiota dysbiosis and barrier dysfunction in inflammatory bowel disease and colorectal cancers: exploring a common ground hypothesis. J Biomed Sci 2018;25:79.

47 Taverniti V, Stuknyte M, Minuzzo M, Arioli S, De Noni I, Scabiosi C, et al. S-layer protein mediates the stimulatory effect of Lactobacillus helveticus MIMLh5 on innate immunity. Applied and environmental microbiology 2013;79:1221–31.

48 Zheng D, Liwinski T, Elinav E. Interaction between microbiota and immunity in health and disease. Cell Res 2020;30:492–506.

49 Wasinger VC, Lu K, Yau YY, Nash J, Lee J, Chang J, et al. Spp24 is associated with endocytic signalling, lipid metabolism, and discrimination of tissue integrity for ‘leaky-gut’ in inflammatory bowel disease. Sci Rep 2020;10:12932.

50 Chelakkot C, Ghim J, Ryu SH. Mechanisms regulating intestinal barrier integrity and its pathological implications. Exp Mol Med 2018;50:1–9.

51 Bertocchi A, Carloni S, Ravenda PS, Bertalot G, Spadoni I, Lo Cascio A, et al. Gut vascular barrier impairment leads to intestinal bacteria dissemination and colorectal cancer metastasis to liver. Cancer Cell 2021;39:708–24 e11.

52 Wei EK, Giovannucci E, Wu K, Rosner B, Fuchs CS, Willett WC, et al. Comparison of risk factors for colon and rectal cancer. Int J Cancer 2004;108:433–42.

53 Paschke S, Jafarov S, Staib L, Kreuser ED, Maulbecker-Armstrong C, Roitman M, et al. Are Colon and Rectal Cancer Two Different Tumor Entities? A Proposal to Abandon the Term Colorectal Cancer. Int J Mol Sci 2018;19.

54 Zhang J, Haines C, Watson AJM, Hart AR, Platt MJ, Pardoll DM, et al. Oral antibiotic use and risk of colorectal cancer in the United Kingdom, 1989-2012: a matched case-control study. Gut 2019;68:1971–8.

55 Gaertner WB, Kwaan MR, Madoff RD, Melton GB. Rectal cancer: An evidence-based update for primary care providers. World J Gastroenterol 2015;21:7659–71.

56 Hjartaker A, Aagnes B, Robsahm TE, Langseth H, Bray F, Larsen IK. Subsite-specific dietary risk factors for colorectal cancer: a review of cohort studies. J Oncol 2013;2013:703854.

57 Rossi M, Mascaretti F, Parpinel M, Serraino D, Crispo A, Celentano E, et al. Dietary intake of branched-chain amino acids and colorectal cancer risk. Br J Nutr 2021;126:22–7.

58 Lee GH, Malietzis G, Askari A, Bernardo D, Al-Hassi HO, Clark SK. Is right-sided colon cancer different to left-sided colorectal cancer? - a systematic review. Eur J Surg Oncol 2015;41:300–8.

59 Glebov OK, Rodriguez LM, Nakahara K, Jenkins J, Cliatt J, Humbyrd CJ, et al. Distinguishing right from left colon by the pattern of gene expression. Cancer Epidemiol Biomarkers Prev 2003;12:755–62.

60 Tamas K, Walenkamp AM, de Vries EG, van Vugt MA, Beets-Tan RG, van Etten B, et al. Rectal and colon cancer: Not just a different anatomic site. Cancer Treat Rev 2015;41:671–9.

61 Mima K, Cao Y, Chan AT, Qian ZR, Nowak JA, Masugi Y, et al. Fusobacterium nucleatum in Colorectal Carcinoma Tissue According to Tumor Location. Clin Transl Gastroenterol 2016;7:e200.

62 Bergstrom K, Shan X, Casero D, Batushansky A, Lagishetty V, Jacobs JP, et al. Proximal colon-derived O-glycosylated mucus encapsulates and modulates the microbiota. Science 2020;370:467–72.

63 Ransohoff DF. Evaluating a New Cancer Screening Blood Test: Unintended Consequences and the Need for Clarity in Policy Making. J Natl Cancer Inst 2021;113:109–11.

64 Allison JE, Tekawa IS, Ransom LJ, Adrain AL. A comparison of fecal occult-blood tests for colorectal-cancer screening. N Engl J Med 1996;334:155–9.

65 Boleij A, Tjalsma H. The itinerary of Streptococcus gallolyticus infection in patients with colonic malignant disease. The Lancet Infectious diseases 2013;13:719–24.

66 Wang HF, Li LF, Guo SH, Zeng QY, Ning F, Liu WL, et al. Evaluation of antibody level against Fusobacterium nucleatum in the serological diagnosis of colorectal cancer. Sci Rep-Uk 2016;6:33440.

67 Franco DL, Leighton JA, Gurudu SR. Approach to Incomplete Colonoscopy: New Techniques and Technologies. Gastroenterol Hepatol (N Y) 2017;13:476–83.

