## Supplemental material for "Blood bacterial DNA, intestinal adenoma and colorectal cancer"

SUPPLEMENTARY MATERIAL

### Supplementary Methods

*Bacterial DNA contamination assessment*

To assess the potential bacterial DNA contamination from environment and reagents, several negative controls were included and analysed throughout the 16S rRNA gene sequencing pipeline. Ten negative controls consisting of ultrapure water (NC-EXT-WA) were added at the DNA extraction step to account for reagent and environmental contaminants. NC-EXT-WA samples were processed concomitantly with the DNA extraction from whole blood samples. This study comprised 296 whole blood samples. 16 rounds of DNA extraction were performed adding 2 NC-EXT-WA samples to the extraction round 1 (EXT1), 5 (EXT5), 10 (EXT10), 15 (EXT15) and 16 (EXT16).

To overcome the unbalance between the whole blood sample and the control groups (296 vs 10), we performed 10 randomized statistical analyses by comparing the 10 NC-EXT-WA samples to 10 randomly selected whole blood samples. The beta-diversity was measured through Bray-Curtis index. PERMANOVA and PERMDISP were used to test for homogeneity of multivariate dispersions on Bray-Curtis dissimilarity table for the contamination control.

We used diversity analyses and comparisons of the taxonomic profiles in order to confirm that the differences between DNA extraction negative controls and whole blood samples were large enough. Beta-diversity indices were used to investigate whether blood bacterial DNA differs from that of extraction negative controls. Based on all beta-diversity indices, especially on the Bray-Curtis distance matrix (Supplementary Figure 4), blood bacterial DNA profiles were separated from the negative controls in all the randomized analyses. Moreover, blood samples tended to form a tighter cluster than the negative controls, suggesting that the blood samples shared a specific and more similar DNA bacterial profile. The negative control group was more dispersed, showing more variability in-between samples and suggesting a random profile more evident. PERMANOVA showed statistically significant differences between whole blood samples and negative controls for Bray-Curtis beta-diversity indices (Supplementary Table 2), suggesting that the bacterial communities differed both in composition and relative abundance between biological and negative control samples. PERMDISP confirmed the results.

We could, therefore, consider that the background noise did not impact the results of the study.

*Supplementary Table 1* Distributions of Observed, Chao1, Shannon and Simpson alpha-diversity indices of control, intestinal adenoma (IA), colon and rectal cancer subjects for bacterial genera and OTUs.

*Supplementary Table 2:* PERMANOVA and PERMDISP on beta-diversity Bray-Curtis index for the 10 randomized comparisons between negative controls (CTRL) and whole blood samples (WB). For all randomized comparisons, the sample size is equal to 20 (10 Neg CTRL vs. 10 WB) at the exception of RandExt16 (10 Neg CTRL vs. 9 WB). The statistical testing used were pseudo-F and F-value for PERMANOVA and PERMDISP, respectively. For both tests, 2000 permutations were performed. P-values<0.05 in yellow, <0.005 in orange and <0.0005 in red.

*Supplementary Figure 1.* Distribution of 16S rRNA gene copies per µl of whole blood among controls, intestinal adenomas and colorectal cancers.

*Supplementary Figure 2.* Mean of the taxonomy composition in terms of relative abundance per group of samples at family taxonomic level. The panel on the left shows the taxonomic composition of controls, intestinal adenoma subjects (IA) and both controls/IA, respectively (as per legend to the right side). The panel on the right shows the taxonomic composition of colorectal cancer (CRC), and rectal and colon cancer cases separately (as per legend to the right side).

*Supplementary Figure 3.* Different taxa between: (A) colorectal cancers (CRC) and controls; (B) CRC and intestinal adenoma (IA); (C) IA and controls by DESeq2 analyses. The taxonomic lineage of each taxon is shown: p, phylum; c, class; o, order; f, family; g, genus; OTU#, Operational Taxonomic Unit. The first two columns show the logarithmic transformation of normalized base mean value for each group. The "padj" column shows the p-value for heterogeneity between groups adjusted for multi-testing analyses. Positive fold changes (shown on a green background) designate taxon overrepresentation in the CRC group. Negative fold changes (shown on a yellow background) designate taxon underrepresentation in the CRC group.

*Supplementary Figure 4.* Multidimensional scaling (MDS) differentiates bacterial patterns of study blood samples and DNA extraction negative controls on Bray-Curtis distance matrices for all 10 randomized comparisons. Study blood samples (in Red) separated from negative controls (in Blue) in all of the randomized comparisons.

**Supplementary Table 1** Distributions of Observed, Chao1, Shannon and Simpson alpha-diversity indices of control, intestinal adenoma (IA), colon and rectal cancer subjects for bacterial genera and OTUs.

| | | Median (I-III quartiles) | | | | p* | p* | p* | p* | p* | p* |
|---|---|---|---|---|---|---|---|---|---|---|---|
| | *Alpha-diversity* | Controls | IA | Colon cancer | Rectal cancer | Controls vs IA | Colon cancer vs IA | Colon cancer vs controls | Rectal cancer vs IA | Rectal cancer vs controls | Colon cancer vs rectal cancer |
| **Genera** | *Observed* | 28.5 (25-32) | 28 (25-32) | 29.5 (25-36) | 30 (26-32) | 0.917 | 0.246 | 0.251 | 0.364 | 0.332 | 0691 |
| | *Chao1* | 44.9 (33-57.5) | 44.5 (34-55.5) | 44.5 (36.3-55.3) | 41.3 (35-53) | 0.686 | 0.874 | 0.774 | 0.617 | 0.879 | 0.493 |
| | *Shannon* | 2.34 (2.02-2.53) | 2.33 (2.15-2.51) | 2.33 (2.00-2.58) | 2.32 (2.12-2.51) | 0.575 | 0.824 | 0.706 | 0.958 | 0.540 | 0.723 |
| | *Simpson* | 0.87 (0.80-0.89) | 0.86 (0.83-0.89) | 0.86 (0.81-0.90) | 0.86 (0.82-0.89) | 0.843 | 0.808 | 0.884 | 0.855 | 0.792 | 0.981 |
| **OTUs** | *Observed* | 34 (31-39) | 35 (31-42) | 37 (32-47) | 37 (33-40) | 0.542 | 0.208 | 0.105 | 0.440 | 0.171 | 0.617 |
| | *Chao1* | 62.1 (48-85) | 66 (49.2-90.5) | 67.1 (51.0-87.5) | 59.9 (49.6-82) | 0.297 | 0.631 | 0.594 | 0.299 | 0.968 | 0.702 |
| | *Shannon* | 2.58 (2.28-2.77) | 2.58 (2.40-2.80) | 2.61 (2.27-2.87) | 2.51 (2.41-2.74) | 0.372 | 0.874 | 0.496 | 0.625 | 0.667 | 0.581 |
| | *Simpson* | 0.90 (0.86-0.92) | 0.90 (0.87-0.92) | 0.90 (0.86-0.92) | 0.89 (0.88-0.91) | 0.671 | 0.879 | 0.749 | 0.241 | 0.482 | 0.378 |

* P for heterogeneity estimated from the Wilcoxon rank-sum
SD: standard deviation; IQ: interquartile

***Supplementary Table 2:*** *PERMANOVA and PERMDISP on beta-diversity Bray-Curtis index for the 10 randomized comparisons between negative controls (CTRL) and whole blood samples (WB). For all randomized comparisons, the sample size is equal to 20 (10 Neg CTRL vs. 10 WB) at the exception of RandExt16 (10 Neg CTRL vs. 9 WB). The statistical testing used were pseudo-F and F-value for PERMANOVA and PERMDISP, respectively. For both tests, 2000 permutations were performed. P-values<0.05 in yellow, <0.005 in orange and <0.0005 in red.*

| | | | RandExt01 | RandExt05 | RandExt10 | RandExt15 | RandExt16 | RandAllExt | Rand1 | Rand2 | Rand3 | Rand4 |
|---|---|---|---|---|---|---|---|---|---|---|---|---|
| Bray-Curtis | PERMANOVA | Test statistic | 3,49 | 4,51 | 3,51 | 4,01 | 4,37 | 3,03 | 4,08 | 3,22 | 2,94 | 2,82 |
| | | P-value | 0,0005 | 0,0005 | 0,0005 | 0,0005 | 0,0005 | 0,0005 | 0,0005 | 0,0005 | 0,0005 | 0,0005 |
| | PERMDISP | Test statistic | 10,67 | 29,79 | 15,52 | 13,87 | 28,65 | 9,96 | 15,13 | 18,59 | 8,28 | 3,60 |
| | | P-value | 0,0050 | 0,0005 | 0,0025 | 0,0005 | 0,0005 | 0,0130 | 0,0005 | 0,0015 | 0,0190 | 0,0900 |

**Supplementary Figure 1.** Distribution of 16S rRNA gene copies per µl of whole blood among controls, intestinal adenomas and colorectal cancers.

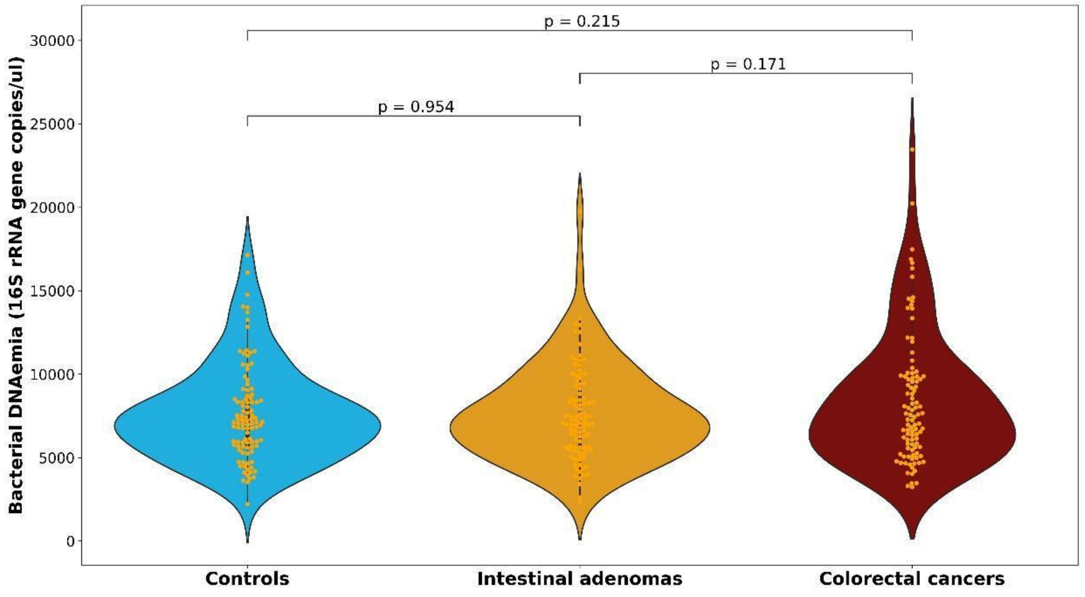

***Supplementary Figure 2.*** Mean of the taxonomy composition in terms of relative abundance per group of samples at family taxonomic level. The panel on the left shows the taxonomic composition of controls, intestinal adenoma subjects (IA) and both controls/IA, respectively (as per legend to the right side). The panel on the right shows the taxonomic composition of colorectal cancer (CRC), and rectal and colon cancer cases separately (as per legend to the right side).

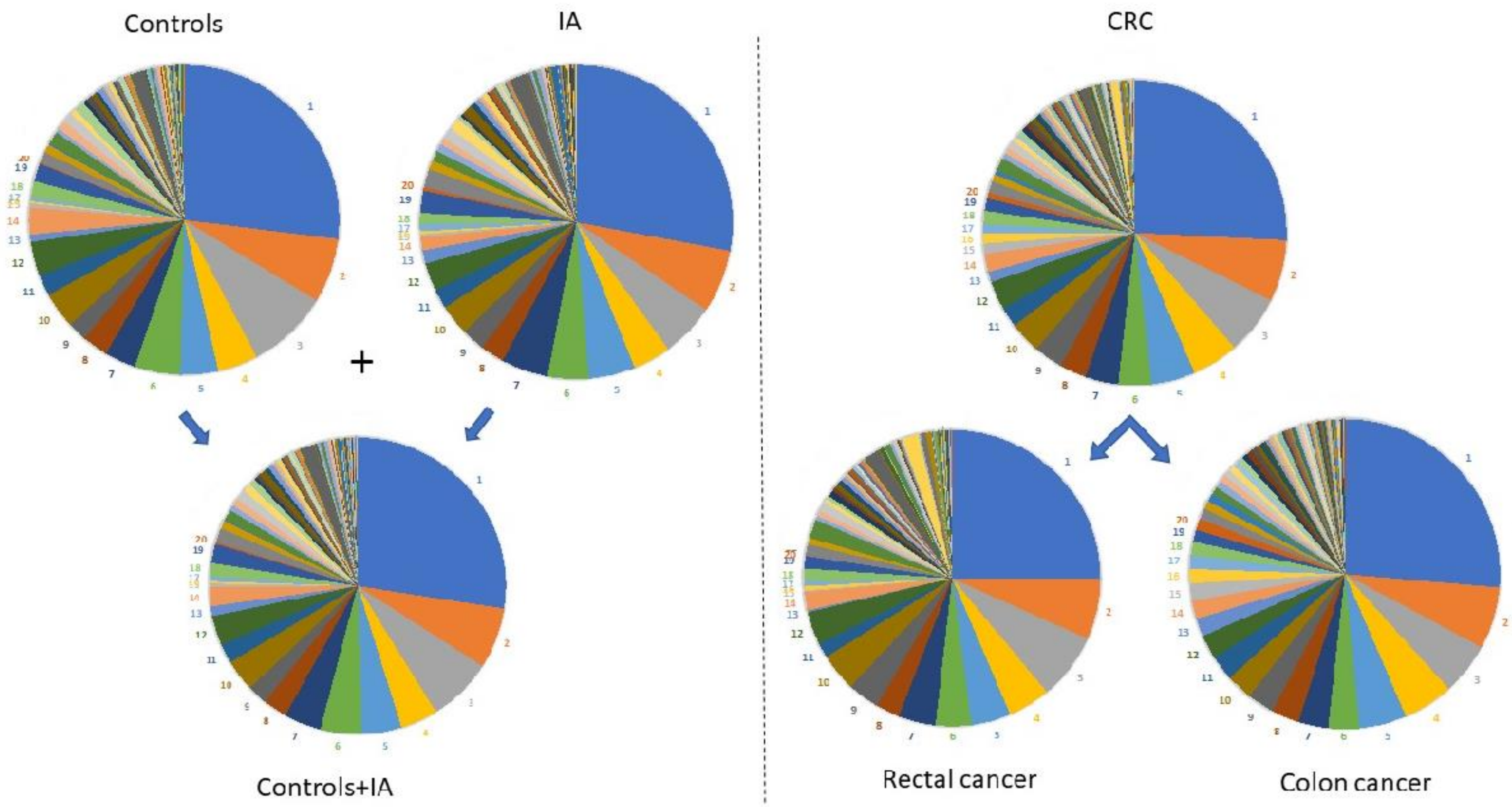


| Number | Family |
|---|---|
| 1 | f_Pseudomonadaceae |
| 2 | f_Micrococcaceae |
| 3 | f_Burkholderiaceae |
| 4 | f_Caulobacteraceae |
| 5 | f_Moraxellaceae |
| 6 | f_Flavobacteriaceae |
| 7 | f_Sphingomonadaceae |
| 8 | f_Legionellaceae |
| 9 | f_Enterobacteriaceae |
| 10 | f_Propionibacteriaceae |
| 11 | f_Corynebacteriaceae |
| 12 | f_Unknown |
| 13 | f_Beijerinckiaceae |
| 14 | f_Xanthomonadaceae |
| 15 | f_Hymenobacteraceae |
| 16 | f_Acetobacteraceae |
| 17 | f_Geodermatophilaceae |
| 18 | f_Rhodobacteraceae |
| 19 | f_Staphylococcaceae |
| 20 | f_Bifidobacteriaceae |

***Supplementary Figure 3.*** Different taxa between: (A) *colorectal cancers (CRC) and controls; (B) CRC and intestinal adenoma (IA); (C) IA and controls* by DESeq2 analyses. The taxonomic lineage of each taxon is shown: p, phylum; c, class; o, order; f, family; g, genus; OTU#, Operational Taxonomic Unit. The first two columns show the logarithmic transformation of normalized base mean value for each group. The "padj" column shows the p-value for heterogeneity between groups adjusted for multi-testing analyses. Positive fold changes (shown on a green background) designate taxon overrepresentation in the CRC group. Negative fold changes (shown on a yellow background) designate taxon underrepresentation in the CRC group.

**A**

| CRC | Controls | #OTU | Taxonomy | padj | log2 Fold Change |
|---|---|---|---|---|---|
| | | | p_Actinobacteria;c_Acidimicrobiia | 3E-02 | 4,92 |
| | | | p_Actinobacteria;c_Actinobacteria;o_Bifidobacteriales;f_Bifidobacteriaceae | 6E-09 | 10,21 |
| | | | p_Actinobacteria;c_Actinobacteria;o_Bifidobacteriales;f_Bifidobacteriaceae;g_Bifidobacterium | 6E-09 | 10,21 |
| | | | p_Actinobacteria;c_Actinobacteria;o_Corynebacteriales;f_Corynebacteriaceae;g_Corynebacterium | 9E-05 | -7,55 |
| | | OTU_54 | p_Actinobacteria;c_Actinobacteria;o_Corynebacteriales;f_Corynebacteriaceae;g_Corynebacterium;s_Corynebacterium sp. | 2E-04 | -7,14 |
| | | | p_Actinobacteria;c_Actinobacteria;o_Frankiales;f_Geodermatophilaceae | 2E-02 | 4,32 |
| | | | p_Actinobacteria;c_Actinobacteria;o_Micrococcales;f_Micrococcaceae;g_Rothia | 8E-08 | 4,45 |
| | | | p_Actinobacteria;c_Actinobacteria;o_o_Bifidobacteriales | 6E-09 | 10,21 |
| | | | p_Actinobacteria;c_Actinobacteria;o_Propionibacteriales;f_Nocardioidaceae | 2E-02 | -4,17 |
| | | | p_Actinobacteria;c_Thermoleophilia | 5E-06 | 7,01 |
| | | | p_Bacteroidetes;c_Bacteroidia;o_Bacteroidales;f_Bacteroidaceae | 2E-09 | -6,77 |
| | | | p_Bacteroidetes;c_Bacteroidia;o_Bacteroidales;f_Bacteroidaceae;g_Bacteroides | 2E-09 | -6,77 |
| | | | p_Bacteroidetes;c_Bacteroidia;o_Sphingobacteriales | 9E-06 | -6,42 |
| | | | p_Bacteroidetes;c_Bacteroidia;o_Sphingobacteriales;f_Sphingobacteriaceae | 1E-04 | -5,39 |
| | | OTU_62 | p_Firmicutes;c_Clostridia;o_Clostridiales;f_Clostridiaceae 1;g_Clostridium sensu stricto 1;s_Multi-affiliation | 9E-05 | -5,04 |
| | | | p_Firmicutes;c_Clostridia;o_Clostridiales;f_Peptostreptococcaceae | 3E-02 | 4,25 |
| | | OTU_49 | p_Firmicutes;c_Clostridia;o_Clostridiales;f_Peptostreptococcaceae;g_Romboutsia;s_metagenome | 1E-06 | 9,20 |
| | | | p_Proteobacteria;c_Alphaproteobacteria;o_Acetobacterales | 5E-06 | 7,38 |
| | | | p_Proteobacteria;c_Alphaproteobacteria;o_Acetobacterales;f_Acetobacteraceae | 5E-06 | 7,38 |
| | | OTU_44 | p_Proteobacteria;c_Alphaproteobacteria;o_Caulobacterales;f_Caulobacteraceae;g_Caulobacter;s_Unknown | 1E-02 | -5,58 |
| | | | p_Proteobacteria;c_Alphaproteobacteria;o_Sphingomonadales;f_Sphingomonadaceae;g_Novosphingobium | 2E-02 | 4,48 |
| | | OTU_42 | p_Proteobacteria;c_Alphaproteobacteria;o_Sphingomonadales;f_Sphingomonadaceae;g_Sphingomonas;s_Multi-affiliation | 9E-05 | -5,34 |
| | | | p_Proteobacteria;c_Deltaproteobacteria;o_Myxococcales | 2E-08 | 4,46 |
| | | | p_Proteobacteria;c_Deltaproteobacteria;o_Oligoflexales;f_Oligoflexaceae | 9E-05 | 7,86 |
| | | OTU_37 | p_Proteobacteria;c_Gammaproteobacteria;o_Betaproteobacteriales;f_Burkholderiaceae;g_Cupriavidus;s_Multi-affiliation | 2E-07 | -7,02 |
| | | | p_Proteobacteria;c_Gammaproteobacteria;o_Betaproteobacteriales;f_Burkholderiaceae;g_Ralstonia | 4E-02 | -4,71 |
| | | | p_Proteobacteria;c_Gammaproteobacteria;o_Betaproteobacteriales;f_Neisseriaceae | 5E-06 | 6,58 |
| | | | p_Proteobacteria;c_Gammaproteobacteria;o_Betaproteobacteriales;f_Neisseriaceae;g_Neisseria | 2E-02 | 4,12 |
| | | | p_Proteobacteria;c_Gammaproteobacteria;o_Enterobacteriales;f_Enterobacteriaceae;g_Pantoea | 2E-04 | 7,73 |
| | | OTU_48 | p_Proteobacteria;c_Gammaproteobacteria;o_Legionellales;f_Legionellaceae;g_Legionella;s_Unknown | 3E-08 | 4,86 |
| | | OTU_83 | p_Proteobacteria;c_Gammaproteobacteria;o_Legionellales;f_Legionellaceae;g_Legionella;s_Unknown | 3E-08 | 5,35 |
| | | OTU_46 | p_Proteobacteria;c_Gammaproteobacteria;o_Pseudomonadales;f_Moraxellaceae;g_Acinetobacter;s_Multi-affiliation | 2E-13 | 10,64 |
| | | OTU_31 | p_Proteobacteria;c_Gammaproteobacteria;o_Pseudomonadales;f_Moraxellaceae;g_Acinetobacter;s_Multi-affiliation | 2E-07 | -7,50 |
| | | OTU_30 | p_Proteobacteria;c_Gammaproteobacteria;o_Pseudomonadales;f_Moraxellaceae;g_Acinetobacter;s_Multi-affiliation | 2E-02 | 4,13 |

**B**

| IA | CRC | #OTU | Taxonomy | padj | log2 Fold Change |
|---|---|---|---|---|---|
| | | OTU_48 | p_Actinobacteria;c_Actinobacteria;o_Corynebacteriales;f_Corynebacteriaceae;g_Corynebacterium 1;s_Unknown | 9E-07 | 7,82 |
| | | OTU_94 | p_Actinobacteria;c_Actinobacteria;o_Micrococcales;f_Microbacteriaceae;g_Microbacterium;s_Microbacterium sp. | 3E-08 | 5,23 |
| | | | p_Actinobacteria;c_Actinobacteria;o_Propionibacteriales;f_f_Nocardioidaceae | 2E-08 | -4,68 |
| | | OTU_6 | p_Actinobacteria;c_Actinobacteria;o_Propionibacteriales;f_Propionibacteriaceae;g_Cutibacterium;s_Multi-affiliation | 7E-08 | 1,16 |
| | | | p_Bacteroidetes;c_Bacteroidia;o_Bacteroidales;f_Bacteroidaceae | 1E-05 | -4,62 |
| | | | p_Bacteroidetes;c_Bacteroidia;o_Bacteroidales;f_Bacteroidaceae;g_Bacteroides | 1E-05 | -4,62 |
| | | OTU_52 | p_Bacteroidetes;c_Bacteroidia;o_Bacteroidales;f_Bacteroidaceae;g_Bacteroides;s_Multi-affiliation | 3E-06 | 6,65 |
| | | | p_Bacteroidetes;c_Bacteroidia;o_Chitinophagales | 1E-06 | 7,47 |
| | | | p_Bacteroidetes;c_Bacteroidia;o_Chitinophagales;f_Chitinophagaceae | 3E-04 | 5,57 |
| | | | p_Bacteroidetes;c_Bacteroidia;o_Cytophagales;f_f_Hymenobacteraceae | 2E-05 | 5,85 |
| | | | p_Bacteroidetes;c_Bacteroidia;o_Cytophagales;f_Hymenobacteraceae;g_Hymenobacter | 5E-04 | 4,86 |
| | | | p_Bacteroidetes;c_Bacteroidia;o_Flavobacteriales;f_Weeksellaceae;g_Cloacibacterium | 4E-05 | -5,64 |
| | | OTU_26 | p_Bacteroidetes;c_Bacteroidia;o_Flavobacteriales;f_Weeksellaceae;g_Cloacibacterium;s_Multi-affiliation | 7E-05 | 5,24 |
| | | | p_Bacteroidetes;c_Bacteroidia;o_Sphingobacteriales | 4E-02 | -3,75 |
| | | | p_Bacteroidetes;c_Bacteroidia;o_Sphingobacteriales;f_Sphingobacteriaceae | 4E-02 | -3,75 |
| | | | p_Dependentiae | 3E-02 | -4,12 |
| | | | p_Dependentiae;c_Babeliae | 3E-02 | -4,12 |
| | | | p_Dependentiae;c_Babeliae;o_Babeliales | 3E-02 | -4,12 |
| | | | p_Firmicutes;c_Bacilli;o_Bacillales;f_Family XI | 9E-07 | 6,90 |
| | | | p_Firmicutes;c_Bacilli;o_Lactobacillales;f_Lactobacillaceae | 2E-07 | 7,56 |
| | | | p_Firmicutes;c_Bacilli;o_Lactobacillales;f_Lactobacillaceae;g_Lactobacillus | 1E-07 | 7,70 |
| | | | p_Firmicutes;c_Clostridia;o_Clostridiales;f_Clostridiaceae;g_Clostridium sensu stricto 1 | 3E-09 | 7,93 |
| | | | p_Firmicutes;c_Clostridia;o_Clostridiales;f_Lachnospiraceae | 4E-02 | -4,01 |
| | | | p_Firmicutes;c_Clostridia;o_Clostridiales;f_Peptostreptococcaceae | 5E-12 | 9,59 |
| | | | p_Firmicutes;c_Clostridia;o_Clostridiales;f_Peptostreptococcaceae;g_Romboutsia | 7E-04 | 5,89 |
| | | OTU_49 | p_Firmicutes;c_Clostridia;o_Clostridiales;f_Peptostreptococcaceae;g_Romboutsia;s_metagenome | 7E-07 | 8,36 |
| | | | p_Firmicutes;c_Clostridia;o_Clostridiales;f_Ruminococcaceae | 2E-08 | -5,41 |
| | | | p_Patescibacteria | 2E-08 | 4,76 |
| | | | p_Patescibacteria;c_Saccharimonadia | 8E-07 | 8,13 |
| | | | p_Patescibacteria;c_Saccharimonadia;o_Saccharimonadales | 8E-07 | 8,13 |
| | | | p_Proteobacteria;c_Alphaproteobacteria;o_Acetobacterales | 4E-08 | 4,70 |
| | | | p_Proteobacteria;c_Alphaproteobacteria;o_Acetobacterales;f_Acetobacteraceae | 4E-08 | 4,70 |
| | | OTU_5 | p_Proteobacteria;c_Alphaproteobacteria;o_Caulobacterales;f_Caulobacteraceae;g_Unknown;s_Unknown | 2E-02 | 1,27 |
| | | OTU_14 | p_Proteobacteria;c_Alphaproteobacteria;o_Rhizobiales;f_Rhizobiaceae;g_Phyllobacterium;s_Multi-affiliation | 3E-02 | -1,71 |
| | | OTU_33 | p_Proteobacteria;c_Alphaproteobacteria;o_Rhodobacterales;f_Rhodobacteraceae;g_Paracoccus;s_Multi-affiliation | 1E-06 | 6,28 |
| | | OTU_42 | p_Proteobacteria;c_Alphaproteobacteria;o_Sphingomonadales;f_Sphingomonadaceae;g_Sphingomonas;s_Multi-affiliation | 9E-04 | -5,01 |
| | | OTU_37 | p_Proteobacteria;c_Gammaproteobacteria;o_Betaproteobacteriales;f_Burkholderiaceae;g_Cupriavidus;s_Multi-affiliation | 4E-05 | 5,77 |
| | | | p_Proteobacteria;c_Gammaproteobacteria;o_Betaproteobacteriales;f_Burkholderiaceae;g_Polynucleobacter | 2E-02 | -6,00 |
| | | OTU_51 | p_Proteobacteria;c_Gammaproteobacteria;o_Betaproteobacteriales;f_Burkholderiaceae;g_Ralstonia;s_Multi-affiliation | 8E-08 | 4,77 |
| | | | p_Proteobacteria;c_Gammaproteobacteria;o_Betaproteobacteriales;f_Neisseriaceae;g_Neisseria | 1E-02 | 4,48 |
| | | | p_Proteobacteria;c_Gammaproteobacteria;o_Enterobacteriales;f_Enterobacteriaceae;g_Pantoea | 4E-05 | 8,05 |
| | | OTU_32 | p_Proteobacteria;c_Gammaproteobacteria;o_Legionellales;f_Legionellaceae;g_Legionella;s_Multi-affiliation | 1E-04 | 5,74 |
| | | OTU_46 | p_Proteobacteria;c_Gammaproteobacteria;o_Pseudomonadales;f_Moraxellaceae;g_Acinetobacter;s_Multi-affiliation | 2E-10 | 8,85 |
| | | OTU_31 | p_Proteobacteria;c_Gammaproteobacteria;o_Pseudomonadales;f_Moraxellaceae;g_Acinetobacter;s_Multi-affiliation | 3E-05 | -6,40 |
| | | OTU_30 | p_Proteobacteria;c_Gammaproteobacteria;o_Pseudomonadales;f_Moraxellaceae;g_Acinetobacter;s_Multi-affiliation | 5E-02 | 3,29 |
| | | OTU_18 | p_Proteobacteria;c_Gammaproteobacteria;o_Xanthomonadales;f_Xanthomonadaceae;g_Stenotrophomonas;s_Multi-affiliation | 5E-10 | 4,63 |

**C**

| IA | Controls | #OTU | Taxonomy | padj | log2 Fold Change |
|---|---|---|---|---|---|
| | | | p_Actinobacteria;c_Actinobacteria;o_Bifidobacteriales | 1E-06 | 9,25 |
| | | | p_Actinobacteria;c_Actinobacteria;o_Bifidobacteriales;f_Bifidobacteriaceae | 1E-06 | 9,25 |
| | | | p_Actinobacteria;c_Actinobacteria;o_Corynebacteriales;f_Corynebacteriaceae;g_Corynebacterium | 3E-05 | -8,05 |
| | | OTU_54 | p_Actinobacteria;c_Actinobacteria;o_Corynebacteriales;f_Corynebacteriaceae;g_Corynebacterium;s_Corynebacterium sp. | 3E-04 | -7,52 |
| | | OTU_48 | p_Actinobacteria;c_Actinobacteria;o_Corynebacteriales;f_Corynebacteriaceae;g_Corynebacterium 1;s_Unknown | 6E-05 | -6,48 |
| | | | p_Actinobacteria;c_Thermoleophilia | 2E-04 | 7,78 |
| | | | p_Bacteroidetes;c_Bacteroidia;o_Chitinophagales | 3E-09 | -8,69 |
| | | | p_Bacteroidetes;c_Bacteroidia;o_Chitinophagales;f_Chitinophagaceae | 6E-09 | -8,83 |
| | | | p_Bacteroidetes;c_Bacteroidia;o_Cytophagales;f_Hymenobacteraceae | 4E-08 | -4,84 |
| | | | p_Bacteroidetes;c_Bacteroidia;o_Cytophagales;f_Hymenobacteraceae;g_Hymenobacter | 4E-08 | -4,85 |
| | | | p_Bacteroidetes;c_Bacteroidia;o_Cytophagales;f_Spirosomaceae | 4E-08 | -5,28 |
| | | | p_Firmicutes;c_Bacilli;o_Lactobacillales;f_Lactobacillaceae | 1E-05 | -6,85 |
| | | | p_Firmicutes;c_Bacilli;o_Lactobacillales;f_Lactobacillaceae;g_Lactobacillus | 8E-06 | -6,99 |
| | | | p_Firmicutes;c_Clostridia;o_Clostridiales;f_Clostridiaceae;g_Clostridium sensu stricto 1 | 5E-13 | -9,39 |
| | | | p_Firmicutes;c_Clostridia;o_Clostridiales;f_Clostridiaceae 1 | 2E-02 | -3,74 |
| | | OTU_62 | p_Firmicutes;c_Clostridia;o_Clostridiales;f_Clostridiaceae 1;g_Clostridium sensu stricto 1;s_Multi-affiliation | 6E-05 | -5,55 |
| | | | p_Firmicutes;c_Clostridia;o_Clostridiales;f_Family XI | 8E-10 | -9,13 |
| | | | p_Patescibacteria | 3E-02 | -3,90 |
| | | | p_Patescibacteria;c_Saccharimonadia | 2E-05 | -7,75 |
| | | | p_Patescibacteria;c_Saccharimonadia;o_Saccharimonadales | 2E-05 | -7,75 |
| | | | p_Proteobacteria;c_Alphaproteobacteria;o_Rhizobiales;f_Beijerinckiaceae;g_Bosea | 3E-02 | -5,05 |
| | | OTU_33 | p_Proteobacteria;c_Alphaproteobacteria;o_Rhodobacterales;f_Rhodobacteraceae;g_Paracoccus;s_Multi-affiliation | 8E-06 | -5,41 |
| | | OTU_37 | p_Proteobacteria;c_Gammaproteobacteria;o_Betaproteobacteriales;f_Burkholderiaceae;g_Cupriavidus;s_Multi-affiliation | 3E-10 | -8,31 |
| | | OTU_71 | p_Proteobacteria;c_Gammaproteobacteria;o_Betaproteobacteriales;f_Burkholderiaceae;g_Massilia;s_Massilia sp. | 1E-06 | 8,81 |
| | | | p_Proteobacteria;c_Gammaproteobacteria;o_Betaproteobacteriales;f_Neisseriaceae | 5E-07 | 7,14 |
| | | OTU_45 | p_Proteobacteria;c_Gammaproteobacteria;o_Diplorickettsiales;f_Diplorickettsiaceae;g_Unknown;s_Unknown | 3E-02 | 4,29 |
| | | OTU_83 | p_Proteobacteria;c_Gammaproteobacteria;o_Legionellales;f_Legionellaceae;g_Legionella;s_Unknown | 2E-02 | 4,36 |
| | | OTU_65 | p_Proteobacteria;c_Gammaproteobacteria;o_Pseudomonadales;f_Moraxellaceae;g_Acinetobacter;s_Unknown | 3E-08 | 10,00 |
| | | OTU_1 | p_Proteobacteria;c_Gammaproteobacteria;o_Pseudomonadales;f_Pseudomonadaceae;g_Pseudomonas;s_Multi-affiliation | 6E-08 | 0,44 |
| | | OTU_18 | p_Proteobacteria;c_Gammaproteobacteria;o_Xanthomonadales;f_Xanthomonadaceae;g_Stenotrophomonas;s_Multi-affiliation | 3E-10 | -4,65 |

-1,3 Base Mean 4

***Supplementary Figure 4.*** Multidimensional scaling (MDS)differentiates bacterial patterns of study blood samples and DNA extraction negative controls on Bray-Curtis distance matrices for all 10 randomized comparisons. Study blood samples (in Red) separated from negative controls (in Blue) in all of the randomized comparisons.

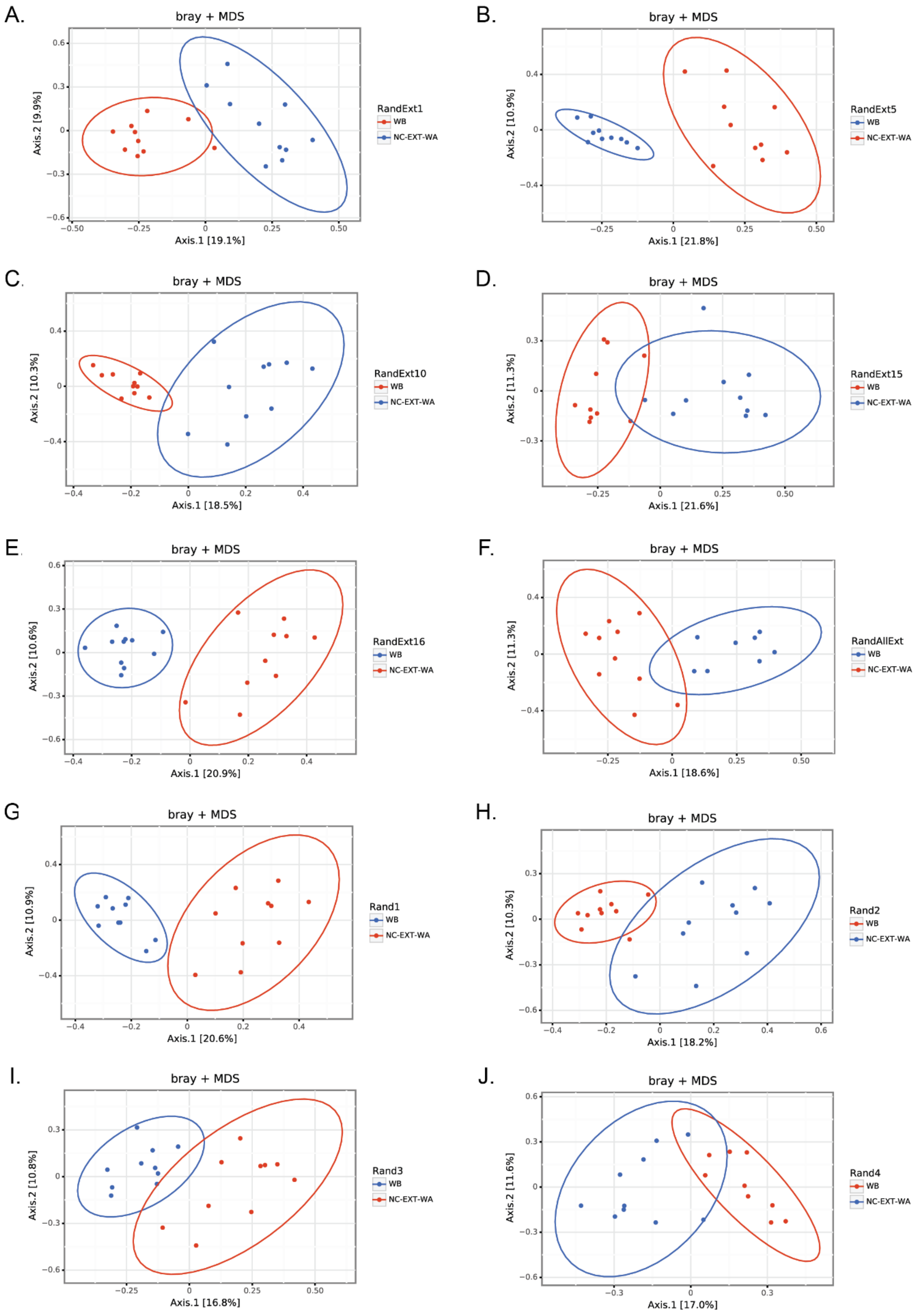
